# Obesity is associated with an altered baseline and post-vaccination influenza antibody repertoire

**DOI:** 10.1101/2021.03.02.21252785

**Authors:** Marwa Abd Alhadi, Lilach M. Friedman, Erik A Karlsson, Liel Cohen-Lavi, Anat Burkovitz, Stacey Schultz-Cherry, Terry L Noah, Samuel S Weir, Lester M. Shulman, Melinda A Beck, Tomer Hertz

**Affiliations:** Department of Microbiology, Immunology and Genetics, Faculty of Health Sciences, Ben-Gurion University of the Negev; Beer-Sheva, Israel; National Center for Biotechnology in the Negev, Ben-Gurion University of the Negev; Beer-Sheva, Israel; Virology Unit, Institute Pasteur du Cambodge; Phnom Penh, Cambodia; Department of Industrial Engineering and Management, Ben-Gurion University of the Negev; Beer-Sheva, Israel; Department of Infectious Diseases, St. Jude Children’s Research Hospital; Memphis, TN, USA; Department of Pediatrics, University of North Carolina at Chapel Hill; NC, USA; Department of Family Medicine, University of North Carolina at Chapel Hill; NC, USA; Dept. of Epidemiology and Preventive Medicine, School of Public Health, Sackler Faculty of Medicine, Tel Aviv University; Tel Aviv, Israel; Department of Nutrition, Gillings School of Global Public Health, UNC, Chapel Hill; NC, USA; Vaccine and Infectious Disease Division, Fred Hutch Cancer Research Center; Seattle, WA, USA

**Author notes:** These authors contributed equally to this work (Marwa Abd Alhadi, Lilach M. Friedman, Melinda A Beck, Tomer Hertz).

## Abstract

As highlighted by the ongoing COVID-19 pandemic, vaccination is critical for infectious disease prevention and control. Obesity is associated with increased morbidity and mortality from respiratory virus infections. While obese individuals respond to influenza vaccination, what is considered a seroprotective response may not fully protect the global obese population. In a cohort vaccinated with the 2010-2011 trivalent inactivated influenza vaccine, baseline immune history and vaccination responses were found to significantly differ in obese individuals compared to healthy controls, especially towards the 2009 pandemic strain of A/H1N1 influenza virus. Young, obese individuals displayed responses skewed towards linear peptides versus conformational antigens, suggesting aberrant obese immune response. Overall, these data have vital implications for the next generation of influenza vaccines, and towards the current SARS-CoV-2 vaccination campaign.

**One Sentence Summary:** Obese individuals have altered baseline and post-vaccination influenza antibody repertoires.

## Main Text

Obesity is an independent risk factor for increased morbidity and mortality after influenza infection, demonstrating the convergence of infectious disease (influenza) with a noncommunicable disease (obesity) (*1*). Similarly, obesity has been shown to be an independent - risk factor for severe outcomes from SARS-CoV-2 infection in the ongoing global COVID-19 pandemic (*2, 3*). The expansive prevalence of obesity worldwide (>500 million obese adults) coupled with significant influenza mortality even in non-pandemic years (3,000-56,000 annual deaths in the US alone) make identifying factors that affect influenza outcomes a critical need. Although vaccination is the primary method of influenza prevention, we have demonstrated that influenza vaccinated adults with obesity have an increased risk of influenza or influenza-like illness compared to vaccinated non-obese adults despite generating what is generally considered a seroprotective response (*4*). To further understand how obesity may impact antibody responses to influenza vaccination, we conducted an in-depth analysis of influenza-specific IgG and IgA antibody repertoires of 205 subjects, 100 with healthy weight (HW: Body mass index (BMI) = 18.5 to 24.9 kg/m2), and 105 with obesity (OB; BMI≥30kg/m2) prior to, and 30 days post-vaccination with split virus trivalent influenza vaccine preparations (TIV; vaccine disrupted by detergent; Table 1). The vaccine was enriched for hemagglutinin (HA) surface protein that contains major conformational neutralizing antigenic epitopes of influenza viruses. The three influenza strains in the 2010-2011 TIV were A/California/7/2009-like virus (A/H1N1; Cal/09), A/Perth/16/2009-like virus (A/H3N2; Perth09) and B/Brisbane/60/2008-like virus (Victoria lineage; BrisB). Of note, this vaccine was the first usage of the Cal09 pandemic strain in the seasonal vaccine, presenting the opportunity to study antibody response to a novel strain (Cal09) and two longer circulating strains (Perth09 and BrisB).

**Table 1.** Demographics of study participants.

| **group** | **N** | **BMI** | **Age** | **Gender**<br>**M/F** | **Race** |
| --- | --- | --- | --- | --- | --- |
| Obese | 105 | 30-46.9 (34.6<br>median) | 20-82.6 ( 54.2<br>median) | 42/63 | Caucasian (55 %)<br>African American (42 %)<br>Hispanic (2 %)<br>Asian (1 %) |
| Healthy-Weight | 100 | 19-24.9 (22.7<br>median) | 19-87 (57.8<br>median) | 41/59 | Caucasian (75 %)<br>African American (16 %)<br>Hispanic (2 %)<br>Asian (6 %) |

Baseline immune history (BIH), the repertoire of antibodies raised against influenza viruses following previous vaccinations and/or natural exposures, varies between individuals. Therefore, we first measured the BIH for IgG and IgA responses to recombinant hemagglutinin proteins (rHA) from Cal09, Perth09, BrisB and 20 additional rHAs obtained from seasonal vaccine and historical strains from 1933-2016 (Table S1) spotted on microarrays. While baseline IgG levels against Cal09 rHA were marginally lower in the OB group (p=0.050; Fig. 1A), baseline IgG levels against rHA antigens of Perth09 and BrisB were similar in OB and HW individuals (Fig. 1B,-C). There was a significantly lower magnitude (sum of antibody levels to all strains from a given subtype) and breadth (number of strains to which a subject has antibodies) for IgG against rHA proteins of A/H1N1 (Fig. 1D, G pre) and lower IgG magnitude to B rHA(Fig. 1F), but not A/H3N2 strains (Fig 2E, H) in OB adults at baseline. There were no differences in BIH IgA responses between HW and OB adults to rHAs of the 3 TIV strains (Fig. S1A-C) or in the BIH IgA magnitude and breadth for A/H3N2 or B proteins in the panel (Fig. S1E-F, H-I). However, the magnitude and breadth of the baseline IgA repertoire against rHA proteins of A/H1N1 strains were significantly decreased in OB individuals, similarly to baseline IgG levels (Fig. S1D, G pre).

**Fig. 1.**
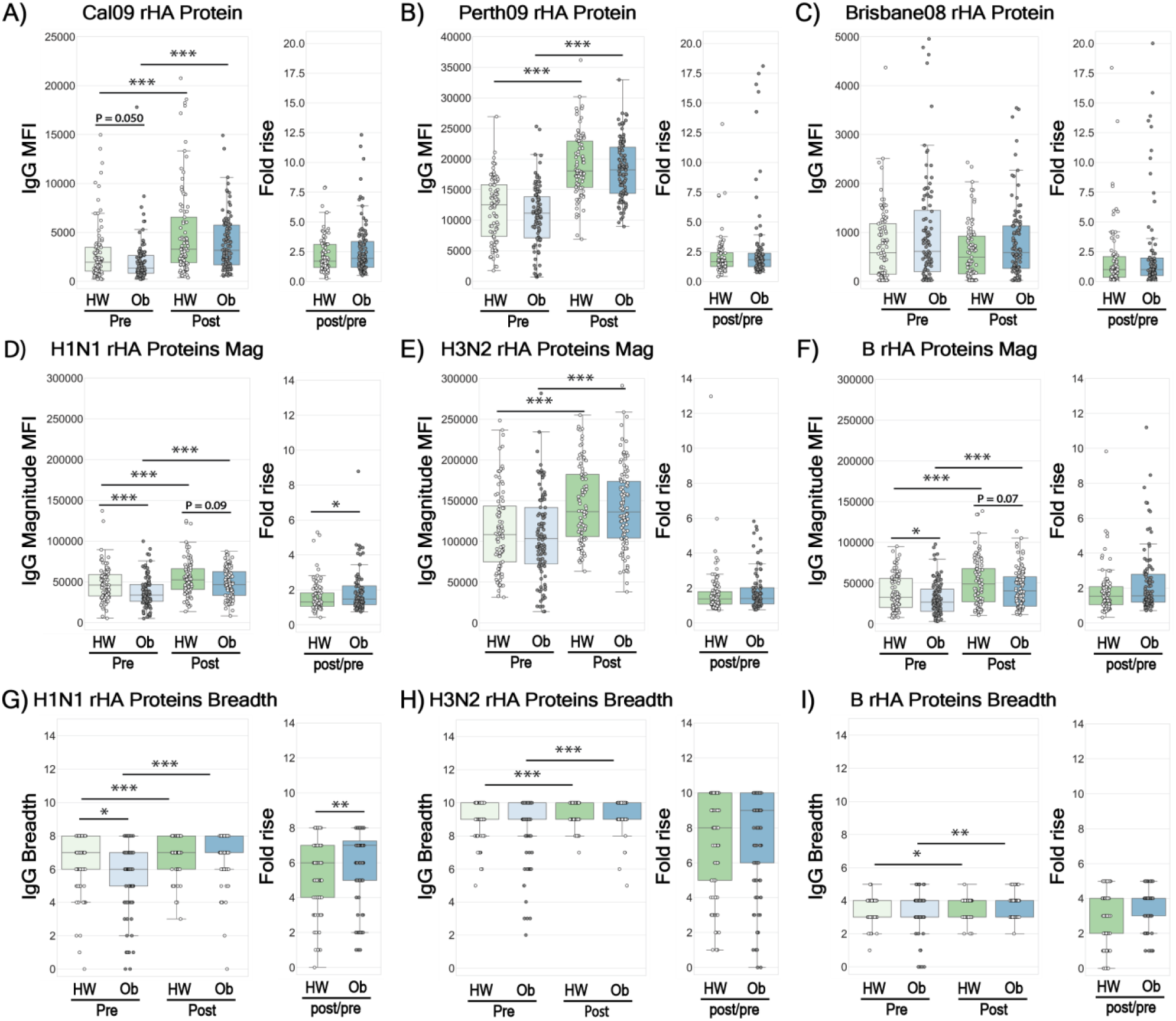
Baseline Immune History (BIH) and 30-day post vaccination IgG responses in healthy weight (HW) and obese (OB) individuals to a panel of historical influenza recombinant proteins (rHA). Baseline and post-vaccination serum samples from 89 healthy-weight (HW) and 100 obese (OB) subjects were hybridized with an antigen microarray spotted with 34 BPL-inactivated influenza viruses and 23 recombinant HA (rHA) proteins that included the three vaccine strains used in the study, for profiling of IgG binding (see Table S1 for IgA binding). **(A-C)** IgG binding to the recombinant HA of H1N1 A/California/7/2009, H3N2 A/Perth/16/2009 and B/Brisbane/60/2008 vaccine strains. (**D-F**) The magnitude of IgG antibodies bound to a panel of 8 recombinant H1 (rH1) proteins (D); a panel of 10 recombinant H3 (rH3) proteins; (E) and a panel of recombinant HA antigens of 5 B strains (F). (**G-I)** The breadth of IgG antibodies bound to a panel of recombinant HA proteins, including: 8 rH1 proteins (G); 10 rH3 proteins (H); and 5 recombinant HA proteins of 5 B strains (l) proteins. The four box plots in the left portion of each panel summarize the baseline (L->R: HW: light green, OB: light blue) and the 30-day post-vaccination (L->R: HW: dark green, OB: dark blue) binding responses. The two boxplots on the right side of each panel represent the fold increase (L->R: HW: green, OB: blue). Lines represent the median fluorescence intensity (MFI), the boxes denote the 25th and 75th percentiles, and the error bars represent 1.5 times the interquartile range. Statistical significance was assessed using the Wilcoxon signed rank test (pre vs. post) and the Wilcoxon rank-sum test (HW vs. OB). * p<0.05, ** p<0.005, *** p<0.0005.

**Fig. 2.**
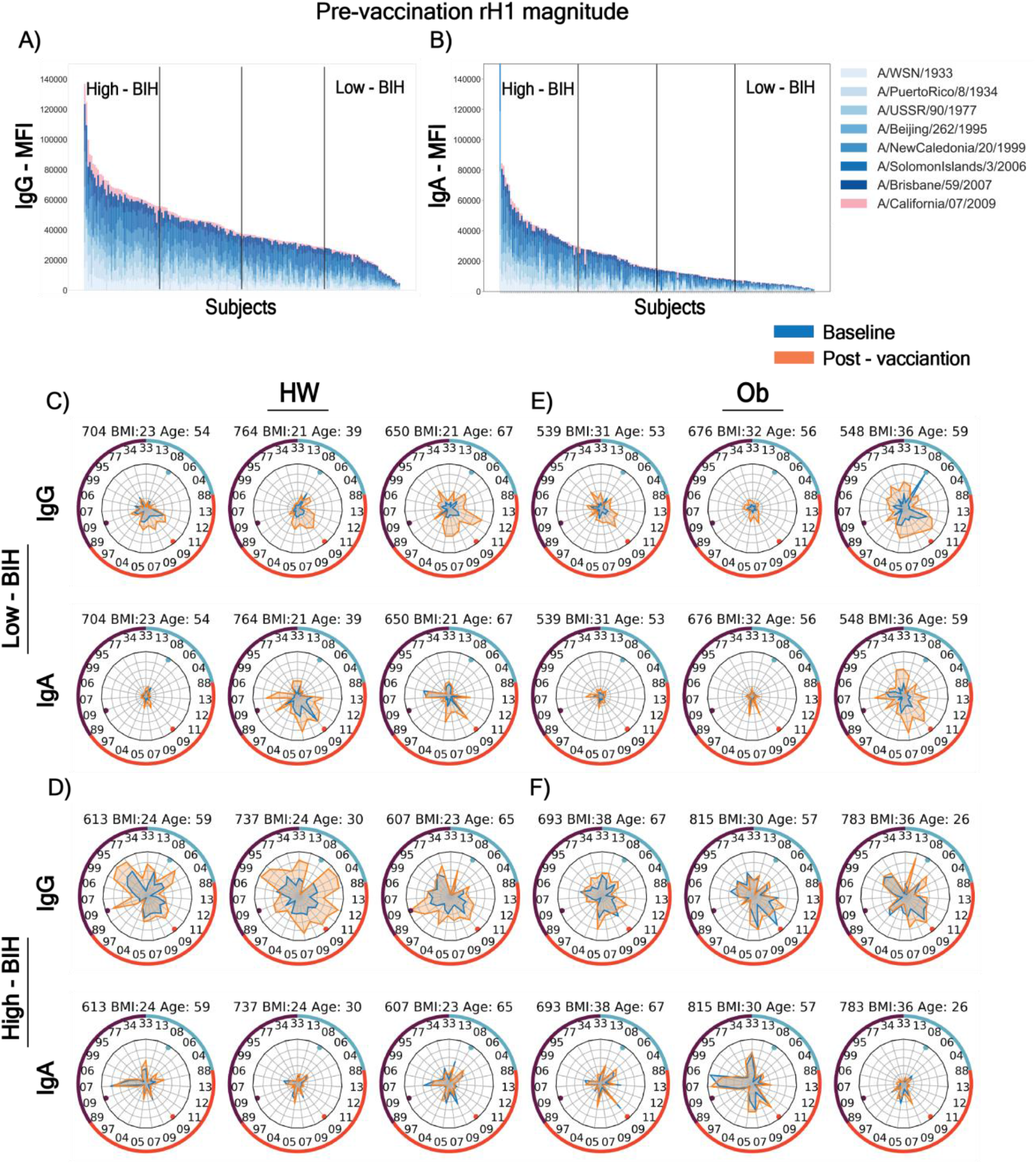
Heterogeneity of baseline (BIH) and 30-day post-vaccination influenza immune history profiles to recombinant HA (rHA) proteins. Panels (A-B): The entire cohort of 205 subjects, 89 healthy-weight (HW) and 100 obese (OB) subjects were assigned to quartiles (low-BIH: n=47, mid-BIH: n=95 and high-BIH: n=47) based on the magnitudes of **(A)** IgG responses and **(B)** IgA responses (low-BIH: n=44, mid-BIH: n=86 and high-BIH: n=44) to recombinant HA proteins of the H1N1 subtype strains. (**C-F**) Spider plots of the BIH and post-vaccination binding responses of individual subjects where each vertex represents the normalized binding of IgG or IgA to a single influenza strain rHA. Baseline and post-vaccination antibody repertoires are denoted by blue and orange spider plots, respectively (light and dark grey in Black and white). The ID, BMI and age of each subject is presented above each spider graph. The numbers on the inside of the outer circles denote the year each strain was isolated, and the three vaccine strains are denoted by the colored dots near the inner circles. Counter-clockwise rHA from: H1N1 (purple segment of circle) strains 33-09-33; H3N2 (red segment of circle) strains 89-13; and B (light blue segment of circle) strains 13-88. Isolates appear in the same order as listed in Table S1. Representative spider graphs are presented for HW subjects (C and D) and OB subjects (E and F) for subjects whose responses were in the low IgG BIH group (C and E) or the high IgG BIH group (D and F).

To further define the specific responses, we measured BIH antibody response against whole, inactivated viruses spotted on microarrays including the 3 TIV strains and a panel of additional 31 whole inactivated influenza viruses (Table S1) representing 83 years of antigenic diversity in A/H1N1, A/H3N2, and B strains between 1933 and 2017. Interestingly, OB individuals had significantly lower total levels of IgG to all 3 TIV strains (Fig. S2A-C), and had significantly decreased IgG magnitude (Fig. S2D-F) and IgG breadth (Fig S2G-I) to all 3 subtypes in the panel. In contrast, there were no significant differences in baseline IgA levels between OB and HW individuals for the current strains (Fig. S3A-C), and no differences in magnitude and breadth of responses to whole viruses (Fig. S3D-I).

Taken together, clear differences in baseline responses exist between OB and HW individuals: the OB group have lower levels of IgG against TIV strains; decreased breadth and magnitude of IgG to 34 strains of whole influenza viruses; and, decreased magnitude and breadth of IgA against rHA proteins from 8 historical A/H1N1 strains of influenza. Thus, prior to vaccination, OB subjects displayed a diminished antibody response against influenza, despite multiple exposures and vaccinations, in particular for subtype A/H1N1.

While OB individuals displayed decreased BIH, previous studies show they do respond to influenza vaccination (*4, 5*). Therefore, to answer whether standard vaccination might overcome obesity-associated BIH deficit in the short term, we measured serum antibody responses at 28-35 days post-vaccination. IgG and IgA levels significantly increased in both HW and OB individuals against rHA of Cal09 and Perth09, but not BrisB, vaccine strains following vaccination (Fig. 1 A-C, S1A-C). Both groups also significantly increased the magnitude and breadth of both IgG and IgA to all three subtypes (Fig. 1D-I; S1D-I). However, vaccine-induced increase in magnitude and breadth of both IgG and IgA against A/H1N1 rHA proteins and IgA against B subtype rHA proteins were significantly stronger in the OB individuals, as reflected by the higher fold-change (Fig. 1 D, G; Fig. S1D, F, G and I). Nevertheless, while the post-vaccination magnitude and breadth to H1N1 rHAs were similar for IgG in both groups (Fig. 1 D, G), they were lower in the OB for IgA (Fig. S1D, G), due to the lower IgA magnitude and breadth at baseline. Post-vaccination increases in breadth or magnitude of IgA or IgG against A/H3N2 or B strains (Fig. S1E-F, H-I, 1E-F, H-I) were not significant between HW and OB individuals.

In terms of whole virus antigens, a significant rise in IgG and IgA levels against all isolates post - vaccination was observed in both OB and HW individuals. However, comparing between groups, total IgG levels and magnitude of response were significantly lower against the three vaccine strains in OB individuals (Fig S2A-F,). In contrast, total IgA levels magnitude and breadth were significantly higher in the OB individuals (Fig. S3D-I). Differences in post - vaccination IgG breadth were noted only for A/H3N2 strains (Fig. S2G-I). In OB individuals, the lower anti-influenza IgG levels post-vaccination were possibly driven by the decreased BIH IgG levels, as the vaccine-induced fold-change in IgG levels, breadth and magnitude were similar between OB and HW (Fig S2). Interestingly, IgA response to the vaccine was stronger in OB individuals reflected by the significantly higher fold-change in IgA to whole influenza viruses, leading to increased IgA levels against whole virus TIV antigens, and higher IgA magnitude and breadth against the three subtypes (Fig S3).

Since antibodies can target linear epitopes in addition to conformational epitopes, we next characterized the BIH and post-vaccination IgG and IgA repertoire using a peptide microarray spotted with a succession of 20-mer peptides with partial overlap of 15 aa spanning the HA and NA protein sequences (H1, N1) of the A/H1N1 Cal09 vaccine strain. This array facilitates characterization of the peptides magnitude, defined as the sum of antibody levels to all peptides from the same protein; peptides breadth, defined as the number of peptides from each protein to which the subject had antibodies; as well as the individual epitopes that were recognized. There was an inverse correlation between recognition of linear epitopes of peptides and recognition of conformational epitopes on whole viruses and rHAs. Specifically, pre- and post-vaccination sera from OB individuals had a higher magnitude and broader repertoire of IgG antibodies to Cal09 H1 and N1 peptides (Fig. S4A-D) in comparison to their lower recognition of Cal09 whole virus and rHA (Fig. 1A, S2A). The pattern for IgAs was more complicated. Pre- and post-vaccination IgA breadth, but not magnitude, to Cal09 H1 and N1 peptides was significantly lower in OB individuals (Fig. S4E-H), while there were no BIH differences for Cal09 whole virus and rHA, but the post-vaccination IgA magnitude and breadth response and fold-responses for H1N1 viruses were greater than for HW individuals (Fig. S3D, G). These findings suggest that an increase in the level of IgG antibodies to conformational antigens is associated with a decrease in the diversity and quantity of IgG antibodies to linear peptides. The differences between the OB and HW antibody response described here were specific to influenza antigens, since the total IgG and IgA levels in the sera of the two groups were similar, as detected by ELISA (Fig. S3J-K).

BIH differs for each individual depending on previous exposure and vaccine history. Indeed, extensive BIH heterogeneity emerged when IgG or IgA baseline magnitude or breadth against specific types of influenza antigens (rHA versus whole viruses versus peptides) was measured in all 205 individuals (e.g. baseline magnitudes of IgG and IgA antibodies to H1N1 rHA (rH1) proteins, Fig. 1D and S1D). To study whether age or obesity affect the BIH to A/H1N1 antigens, the entire cohort was blindly ranked by decreasing BIH IgG and IgA magnitudes of response to whole virus or rHA A/H1N1 antigens, as well as breadth or magnitude of antibodies against Cal09 A/H1 and N1 peptides. For each such ranking, the cohort was divided into quartiles: individuals in the lowest quartile were denoted as low-BIH group, individuals in the highest quartile were denoted as high-BIH group, and the remaining two middle quartiles were denoted as the mid-BIH group (e.g. Fig. 2A-B). To visualize the BIH of each subject, we generated spider plots for a representative subset of low IgG-BIH and high IgG-BIH subjects (Fig. 2C-F), further highlighting that each individual has a unique BIH profile that varies both in the overall magnitude and breadth, but also in the specificity to each subtype and to individual strains within the panel. Interestingly, some subjects with low IgG-BIH to rH1 have low IgG-BIH to all three subtypes (e.g. subjects 704, 539 and 676; Fig. 2C, E), while others have stronger levels of IgG to other subtypes (e.g. subject 548). We found that the antibody response to the vaccine was heavily biased by the individual BIH. While some low-BIH subjects failed to respond to the vaccine (e.g. subject 676), others generated robust vaccine-induced immune responses (e.g. subjects 650 and 548). A comparison between the IgG and IgA spider plots for each subject demonstrated that most subjects with low IgG-BIH to rH1 had also low IgA-BIH to rH1. However, in some cases there were striking differences between IgG and IgA profiles (e.g. subjects 764, 737 and 783, Fig. 2C-F).

The distribution of BMI or age of individuals in the low-BIH and high-BIH groups were compared for each ranking (Fig. 3). This analysis demonstrated again the inverse correlation between magnitude of IgG antibodies to conformational (viruses and proteins) and linear (peptides) antigens when comparing obese and HW groups and when comparing individuals by age. A significantly higher frequency of OB individuals was present in the low-BIH group for magnitude of IgG antibodies to both A/H1N1 viruses (RR 1.86; 95%CI 1.19 to 2.91) and rH1 proteins (RR 2.07; 95%CI 1.29 to 3.34; Fig. 3A-B and S7A). A significantly higher frequency of OB individuals was also present for magnitude of IgA antibodies to rH1 proteins (RR 2.02; 95%CI 1.27 to 3.21), but not to whole H1N1 viruses (RR 1.10; 95%CI 0.72 to 1.69; Fig. 3E-F and S7A). In contrast, when the individuals were ranked according to H1 and N1 peptide response, obese individuals were less frequent in both low-IgG-BIH groups (H1: RR 0.43; 95%CI 0.25 to 0.73; N1: RR 0.43; 95%CI 0.25 to 0.73; Fig. 3E-H, S5A-D andS7A). OB individuals were also more frequently present in the low-BIH group for magnitude of IgG antibodies to A/H3N2 viruses (RR 1.78; 95%CI 1.14 to 2.78), B viruses (RR 1.88; 95%CI 1.18 to 2.98) and B rHI proteins (RR 1.68; 95%CI 1.10 to 2.58) as shown in Fig. S6A, C, D and S7A). Similarly, the age distribution comparison revealed that a significantly higher frequency of individuals younger than 65 was present in the low-BIH group for magnitude of IgG antibodies to A/H1N1 viruses (RR 2.91; 95%CI 1.18 to 7.13), A/H3N2 (RR 2.42; 95%CI 1.10 to 5.32) and B viruses (RR 1.96; 95%CI 1.02to 3.77) as shown in Figure 3I, S6I, S6M and S7B, respectively. This contrasted with lower frequencies of individuals younger than 65 years in the low IgG-BIH breadth groups for H1 (RR 0.60; 95%CI 0.42 to 0.87) and N1 (RR 0.55; 95%CI 0.39 to 0.77) peptides, in Figures 3M, N and S7B).

**Fig. 3.**
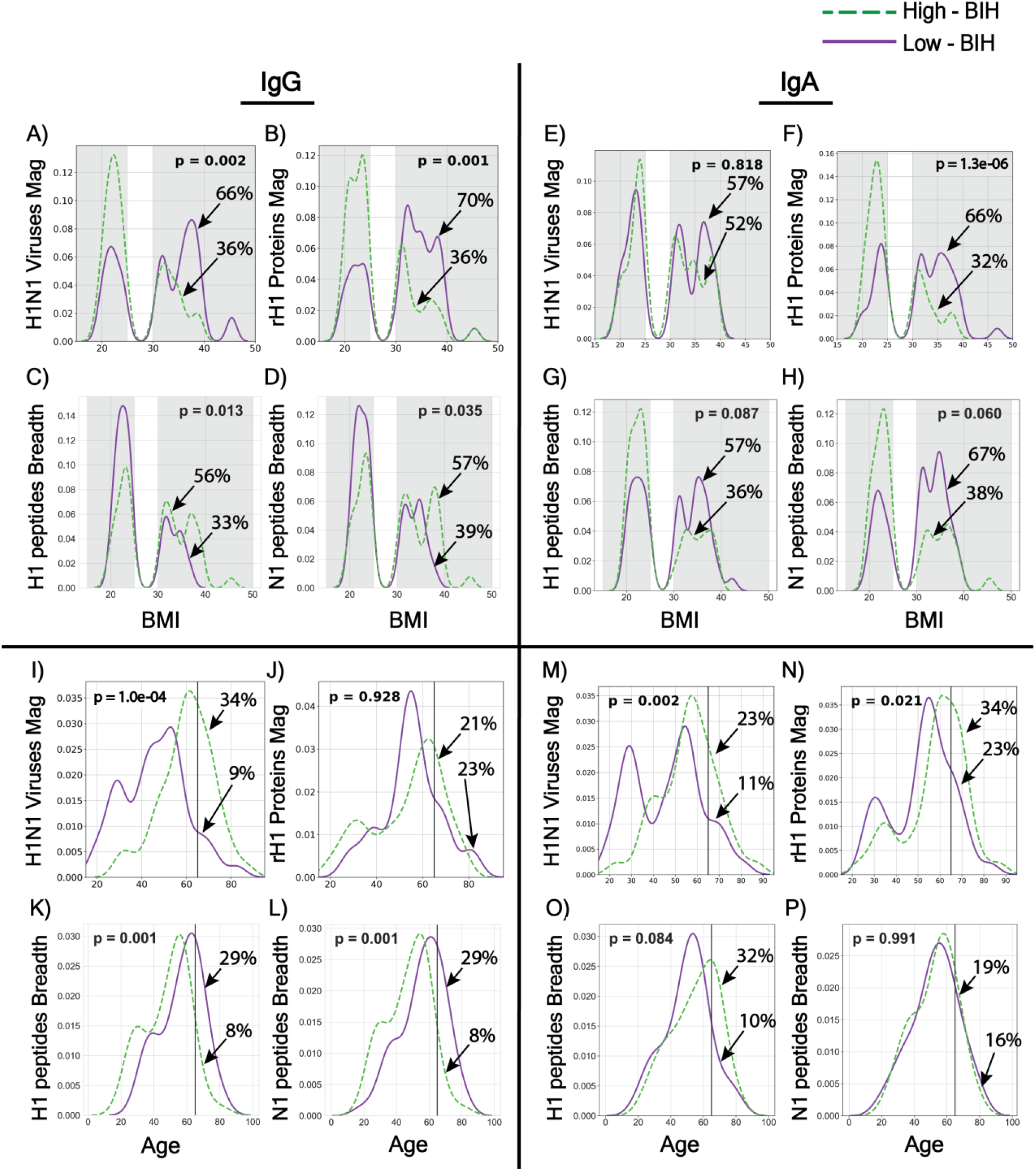
Obesity- and age-associated baseline immune history (BIH) profiles to influenza H1N1 antigens. (**A-H**) The distributions of BMI within the low-BIH group (n=47 for IgG and n=44 for IgA, solid purple line) and the high-BIH group (n=47 for IgG and n=44 for IgA, dashed green line) were plotted for IgG and IgA responses to H1N1 antigens as follows: (A) IgG Magnitude against H1N1 viruses; (B) IgG Magnitude to rH1 proteins; (C) IgG breadth to H1 peptides; (D) IgG breadth to N1 peptides; (E) IgA magnitude to H1N1 viruses; (F) IgA magnitude to rH1 proteins; (G) IgA breadth to H1 peptides; (H) IgA breadth to N1 peptides. Overweight subjects (25 < BMI < 30) were excluded from our analysis. The percentage of obese subjects in the low-BIH responders and high-BIH responders quartiles are listed. (**I- P**): The distributions by subjects age group (<65 y and > 65 y) within the low-BIH group (n=47 for IgG and n=44 for IgA, purple) and the high-BIH group (n=47 for IgG and n=44 for IgA, green) were plotted for their IgG and IgA responses to H1N1 antigens as follows: (I) IgG Magnitude against H1N1 viruses; (J) IgG Magnitude to rH1 proteins; (K) IgG breadth to H1 peptides; (L) IgG breadth to N1 peptides; (M) IgA magnitude to H1N1 viruses; (N) IgA magnitude to rH1 proteins; (O) IgA breadth to H1 peptides; (P) IgA breadth to N1 peptides. p values for differences between the age distributions of the low-BIH and high-BIH groups were determined using the Wilcoxon ranksum test. Obesity was associated with low IgG-BIH to H1N1 viruses and rH1 proteins, high IgA-BIH to rH1 proteins, and high IgG-BIH to peptides of the Cal09 H1 and N1 proteins. Age >65 y was associated with high IgA-BIH to H1N1 viruses and H1 proteins, high IgG-BIH to H1N1 viruses, low IgG- and IgA-BIH to Cal09 H1 peptides, and low IgG-BIH (but not IgA-BIH) to Cal09 N1 peptides. The number of individuals in the low-BIH and high-BIH groups in panels (C-D, G-H, K-L, and O-P) was 47-51 (see Materials and Methods).

The distribution of BMI or age of individuals in the low-BIH and high-BIH groups were then compared for each ranking for IgA responses (Fig. 3 and S6). There were significantly more OB individuals in the low quartile of IgA-BIH to rH1 proteins (Fig. 3F; RR 2.02; 95%CI 1.27 to 3.21, Fig. S7A). Of note, there was a significantly higher number of individuals <65 in the lower IgA-BIH quartiles for IgA responses to magnitude of H1 and N1 peptides (RR 2.18; 95% CI 1.07 to 4.46 for both; Fig. S5G-H and S7B), and there were more individuals <65 in the low quartile of IgA-BIH for H1N1 viruses (Fig 3M), H1N1 proteins (Fig 3N), H3N2 viruses (Fig. S6M), and B viruses (Fig. S6O), although the respective RRs were not significant (1.60, 95% CI 0.76 to 3.38; 1.35, 95% CI 0.79-2.30; 1.60, 95% CI 0.76 to 3.38; and 1.41, 95% CI 0.72 to 2.75 respectively; Fig. S7B). Thus, unlike IgG responses, there was no significant inverse correlation between magnitude of IgA antibodies to conformational (viruses and proteins) and linear (peptides) antigens when taking weight or age into account.

We then checked whether younger and older obese individuals differ by their BIH to H1N1 antigens, since both obese and individuals <65 were enriched in the low-IgG-BIH groups for whole H1N1 viruses, and in the high-IgG-BIH groups for H1 and N1 peptides, as well as in the low-IgA-BIH groups for rH1 proteins (Fig. 3). While no correlation was found between age and BMI (p=0.85), the age distributions of obese individuals in the low-BIH and high-BIH groups were compared. Obese individuals in the low IgG- and IgA-BIH groups to whole viruses were significantly younger than obese subjects in the high-BIH groups to viruses (IgG: p=0.000001 and IgA: p=0.002, respectively). Furthermore, obese individuals in the low-IgA-BIH to rH1 proteins were younger than the obese individuals in the high-IgA-BIH to proteins (p=0.02), while the age of obese individuals in IgG-BIH groups to rH1 proteins was similar (p=0.9). No similar age differences were found when comparing HW individuals from the low-BIH and high-BIH groups.

The significant differences between OB and HW was found in the magnitude and breadth of both IgG and IgA antibodies to Cal09 H1 peptides, prompted to ask whether different peptides were targeted in the OB and HW individuals. We mapped the H1 peptides targeted by BIH IgG and IgA antibodies to the structure of the Cal09 HA protein, and epitopes targeted preferentially by OB and HW individuals were compared using a logistic regression model. Relative weights were assigned to responses to individual peptides, and the score for an individual position was based on the maximal absolute weights assigned to the peptides that overlapped that position. We found that HW IgG antibodies preferentially targeted positions in HA1 subunit and a conserved glycosylation site, while only IgG from OB targeted the more conserved HA2 subunit (Table 2 and Fig. S8D-E). Of the eight functional domains analyzed, five domains (Receptor Binding Site, esterase domain, conserved glycosylation sites, Cal09 glycosylation site and the antigenic sites) were preferentially targeted by IgA of HW individuals, while only the Fusion domain was preferentially targeted by IgA antibodies from OB individuals (Table 2, Fig. 4A-C and S8A-C). These findings further demonstrate that the differences between the HW and OB individuals can be observed even at the resolution of specific functional domains, which may have functional implications for providing protection against infection and development of disease.

**Table 2.** Targeting of HA functional sites by Obese and healthy weight (HW).

|  | pre IgA |  |  |  | pre IgG |  |  |  |
| --- | --- | --- | --- | --- | --- | --- | --- | --- |
|  | Obese | HW | Residues(n) | P Value | Obese | HW | Residues(n) | P Value |
| HA protein | 85 | 114 | 498 |  | 59 | 40 | 498 |  |
| HA1 | 50 | 65 | 322 | 0.7438 | 34 | 40 | 322 | **<1 x 10e-4\*\*** |
| HA2 | 35 | 48 | 174 | 0.3591 | 23 | 0 | 174 | **0.0003\*\*** |
| Fusion | 76 | 53 | 261 | **<1 x 10e-4\*\*** | 33 | 20 | 261 | 0.3008 |
| RBS | 0 | 20 | 153 | **0.0509\*** | 20 | 20 | 153 | 0.9176 |
| Esterase | 7 | 26 | 64 | **0.0006\*\*** | 0 | 0 | 64 | 0.8066 |
| Conserved glycosylation sites | 0 | 6 | 13 | **0.0152\*\*** | 0 | 3 | 13 | **0.0255\*\*** |
| Cal09 glycosylation site | 0 | 3 | 3 | **0.0131\*\*** | 0 | 0 | 3 | 0.8293 |
| Antigenic sites | 0 | 11 | 50 | **0.0371\*\*** | 5 | 0 | 50 | 0.1896 |

\*\* p value below 0.05, \* p value below 0.055

**Fig. 4.**
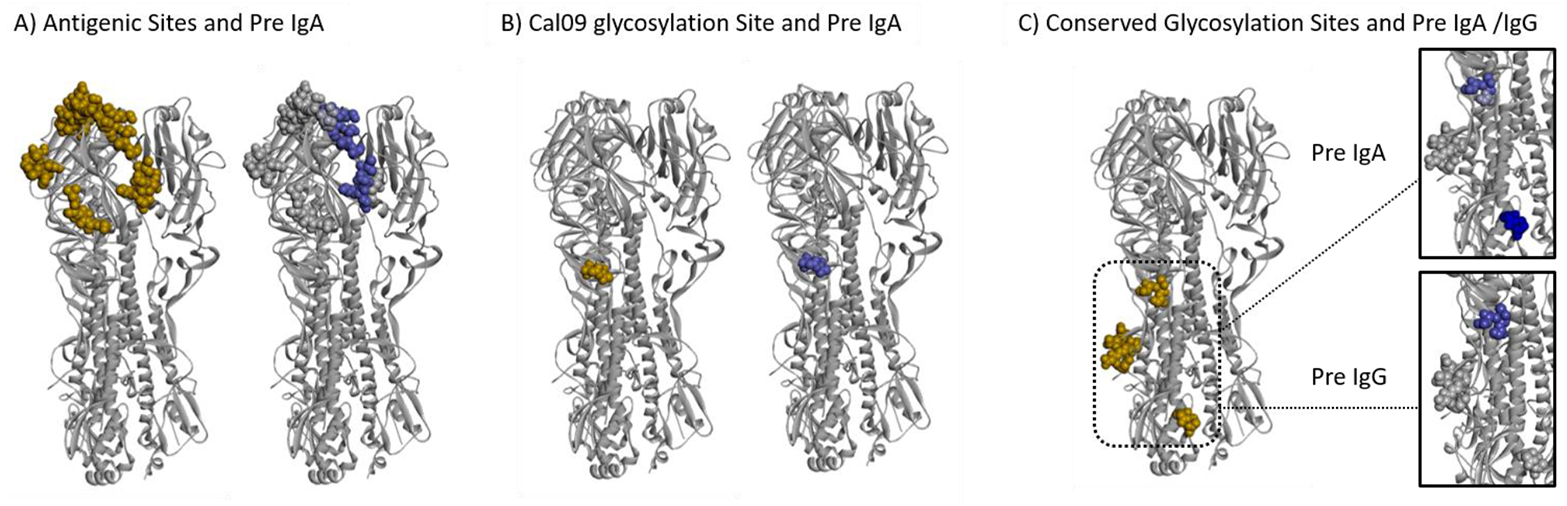
Different domains in the Cal09 HA protein are differentially targeted by BIH antibodies of obese and healthy weight subjects. A logistic regression model was trained to discriminate between HW and OB individuals using the IgG and IgA antibody profiles to the HA peptides of the Cal09 vaccine strain. The weights assigned by the model were used to score individual amino acids on the HA protein based on the maximal weight of a given position across all of the peptides in which it was included (see Materials and Methods for details). Figures were created using Discovery Studio Visualizer software (XXref) and the crystal structure of the Cal09 HA trimeric protein PDB ID: 3LZG (10.1126/science.1186430). Differential binding to additional regions of interest on the HA protein are presented in Figure S9. Left side in each panel: Dark gold spheres represent amino acid residues belonging to three regions of interest mapped onto one of the three trimeric proteins: (**A**) Antigenic sites, (**B**) The Cal09 glycosylation site and (**C**) Conserved Glycosylation sites. Right side in each Panel: Dark Blue spheres represent amino acid residues within the regions of interest associated with antibodies from HW but not OB individuals according to their scores.

Our study population was not controlled for several potential confounders. While age did not appear to differ significantly between HW and OB subjects (Table 1), there were potential asymmetries for gender and race. In addition, other health factors potentially associated with obesity, such as diabetes, smoking history, or hypertension were not accounted for and could potentially have skewed results. For example, 45% of obese subjects and 7% of healthy weight subjects in our cohort suffered from type 2 diabetes. These factors could impact the specificity of our findings for obesity, but would not diminish their potential relevance or vaccine strategies in an immunologically diverse population.

Despite these caveats, this study demonstrates the power of antigen microarrays spotted with different types of antigens presenting both three-dimensional epitopes (whole viruses and rHA proteins) and linear epitopes (overlapping peptides spanning the entire length of proteins being analyzed) for comparing immune responses of different populations to the same vaccine. Consistent with previous studies(*4*), we demonstrate that OB individuals do respond to influenza vaccination; however, this in-depth analysis of the influenza-specific antibody repertoire found striking differences in the antibody repertoires of HW and OB subjects, despite the extensive heterogeneity of the baseline immune-history profiles of subjects in both groups. Adults who were repetitively exposed to influenza viruses by vaccines and infections have a greater opportunity to develop a diverse antibody repertoire; however, antibody responses in OB subjects were suboptimal compared with HW. The comparison of the antibody repertoire against linear and conformational H1N1 antigens in HW and OB subjects demonstrated reduced ability in individuals <65, obese individuals to develop an IgG response to whole virus or rHA protein of the pandemic Cal09 strain, which may be associated with a biased IgG repertoire towards linear epitopes.

Since individuals with obesity were reported as more sensitive to Cal09 infection (*6*), our results suggest that an effective IgG response is associated with IgG antibodies to conformational epitopes. A recent study found that influenza vaccination in humans can recruit memory B cells that were developed following previous exposures to influenza, into the germinal centers, for a further affinity maturation (*7*). We hypothesize that a successful antibody response following exposure of vaccination may convert immature antibodies to linear epitopes to more effective and possibly better neutralizing antibodies against conformational epitopes. Further research of the memory B cells recruited to germinal centers following influenza vaccination is required to test this hypothesis. On the other hand, a broader IgG repertoire against Cal09 H1 and N1 peptides may confer increased cross-reactivity to historical and future pandemic and endemic strains. Additional functional studies are necessary to understand how to harness differential BIH and vaccine-induced immune responses of the obese population in order to induce more effective antibody responses, especially once it is biased toward linear epitopes for IgG. Our data suggest that different influenza vaccines may be required for achieving optimal protection in an obese population.

The OB IgG antibody repertoire appears to be biased towards linear peptides and not whole viruses and rHA proteins, whereas the baseline OB IgA repertoire to H1N1 antigens is less biased. The IgA repertoire is narrower for both rH1 proteins and Cal09 H1 peptides. Taken together, these findings suggest that the IgG and IgA repertoires develop independently, or may be a function of infection during a novel pandemic. It is unclear why obesity affects the IgG repertoire more than the IgA repertoire, and additional mechanistic studies are required to address this question.

While the exact reason for the difference between obese and HW antibody responses to influenza remains to be studied, childhood obesity and even maternal obesity are associated with increased risk of obesity in adulthood (*8-11*). These early obesity factors could lead to aberrant immune response to vaccination and/or infection, leading to differential “original antigenic sin” compared to HW children or adults. Indeed, younger OB individuals in this cohort were more frequent in the lowest quartiles of BIH to H1N1 whole viruses and rHA proteins, which could bias responses later in life, especially if these individuals remain obese. Indeed, the fact that the majority of aberrant responses were observed with the relatively “novel” antigen, the 2009 pandemic strain, indicates that primary or early responses to specific antigens can result in significantly skewed responses. Further longitudinal studies are needed on birth cohorts to determine if childhood obesity or other factors may help to predict vaccination failure later in adulthood, even in HW individuals.

In addition accumulated differences in yearly protection against seasonal human influenza strains, and possibly to vaccines against future emergence of avian influenza strains, this work also has direct and timely relevance to the response to the current global COVID-19 pandemic. Influenza vaccines remain the best way to reliably prevent influenza infections. Due to the speculated cocirculation of SARS-CoV-2 and influenza viruses in the Northern Hemisphere during winter months and the similarity of symptoms between disease manifestations, it is imperative for broad coverage of influenza vaccination to reduce clinical and diagnostic burden as well as morbidity and mortality from influenza virus in an already straining healthcare system. In addition, the understanding of humoral immunity and vaccination effectiveness and duration against SARS-CoV-2 is nascent. As obesity is already considered a high risk factor in disease severity and mortality, if there is a differential response to specific vaccines, even with equivalent overall serological response, OB individuals may still be at risk for contracting and spreading SARS-CoV-2 within the population. Finally, the existence of significant numbers of asymptomatic infections is a confounding factor for predicting potential response of vaccines as these individuals are no longer naïve and obesity may skew initial antibody responses, therefore making future vaccination efforts ineffective, or, more concerningly, driving pathogen evolution and antibody escape. Overall, the design and development of different vaccines for OB individuals may need to be seriously taken into consideration, and clinical trials should take potential differences in BIH and vaccine-induced responses into account when choosing a representative “healthy” study population for any phase.

## Data Availability

All raw data and code used in the analysis are available upon request

## Acknowledgements

The authors thank the Israel Science Foundation grant 882/17 (TH), The Israel America Foundation and the Ben-Gurion University Center for Multidisciplinary Research in Aging (TH), National Institute of Allergy and Infectious Diseases CEIRS contract under HHS contract HHSN27220140006C (EAK and SSC), ALSAC (EAK and SSC), as well as R01 NIH/NIAID/AI078090 (MAB, TLN and SSW) for supporting the study.

## Author contributions

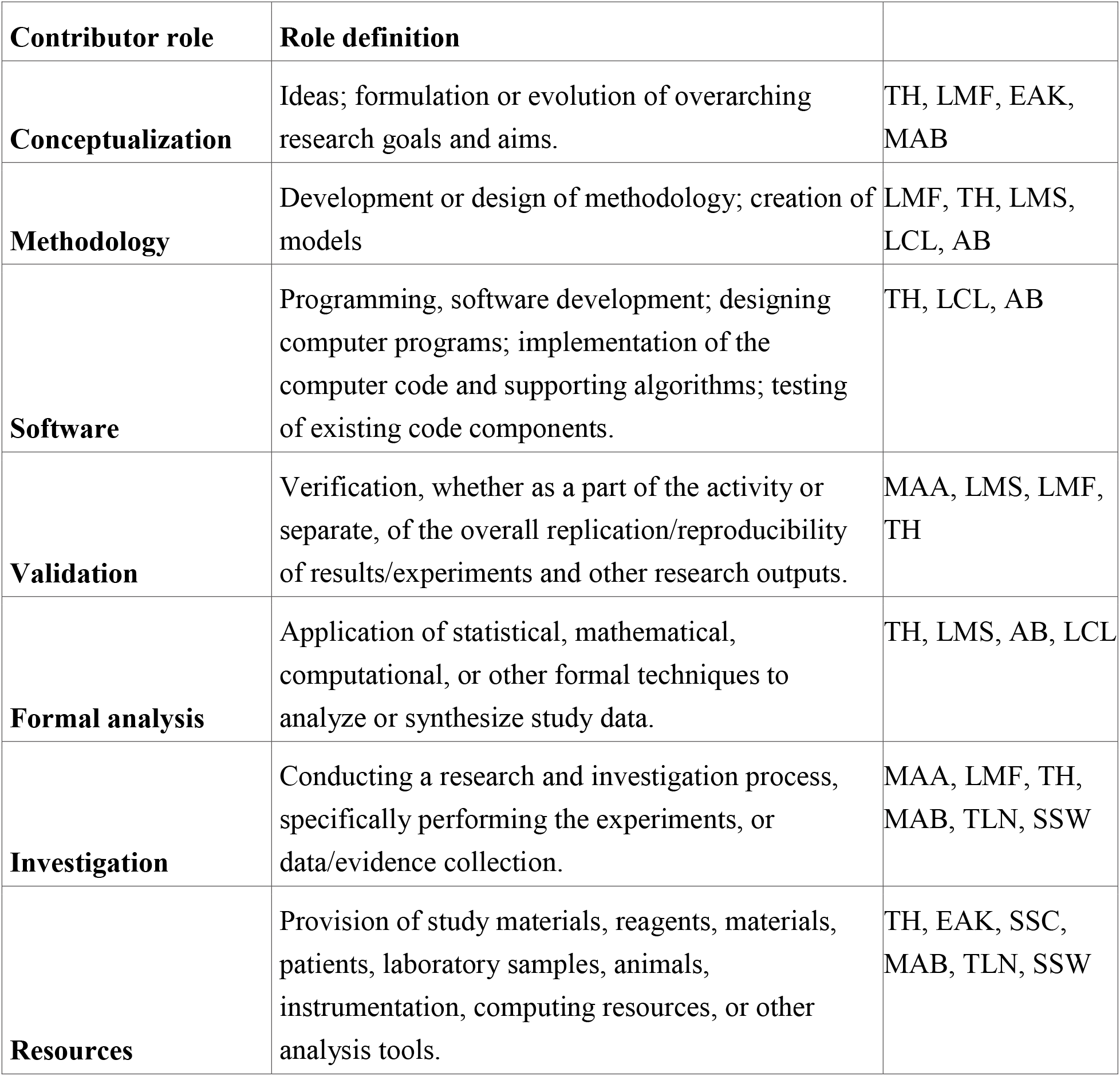
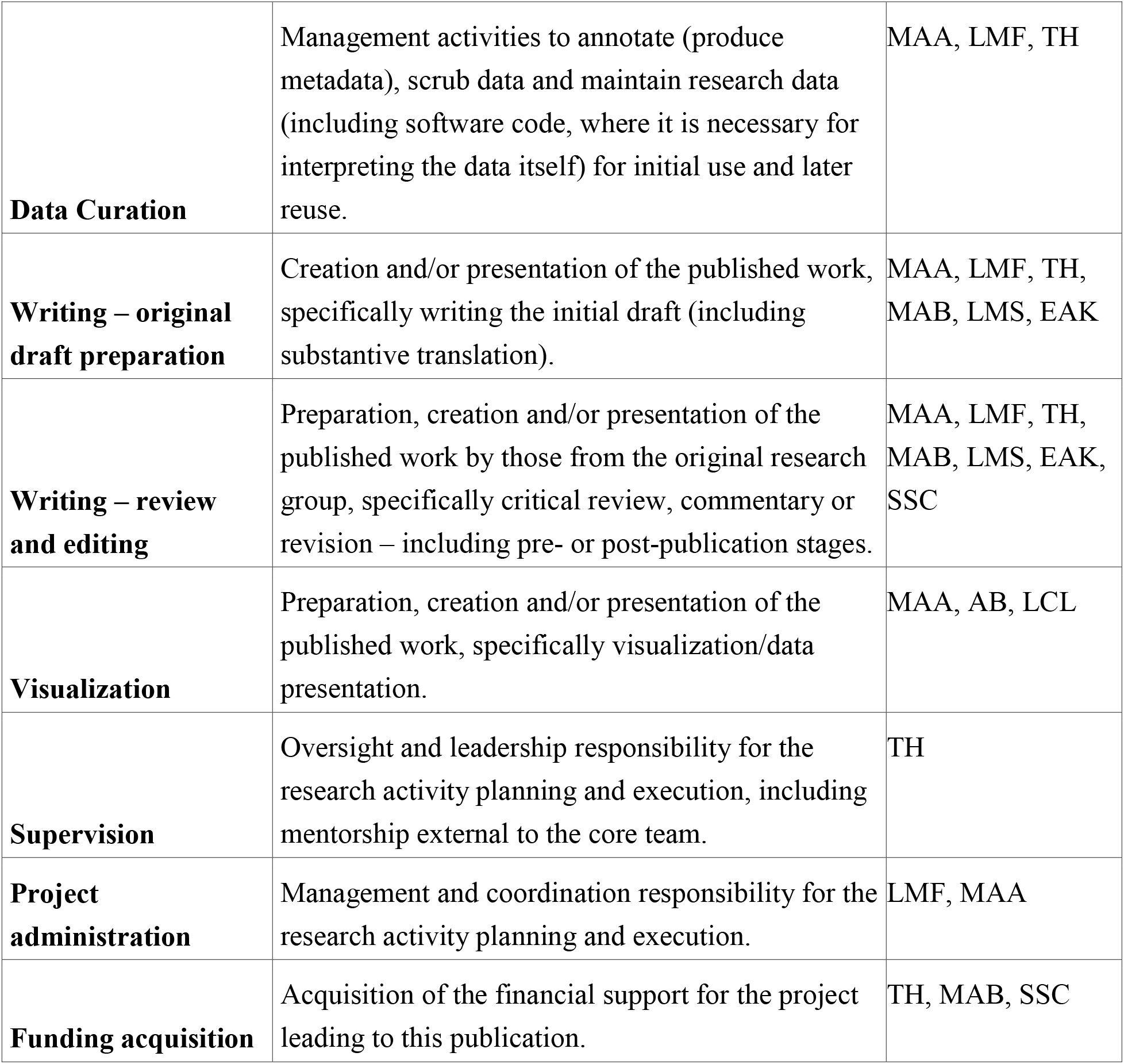

## Data and materials availability

All row data and code used in the analysis are available in the Hertz Lab website: https://www.hertz-lab.org/

## Supplementary Materials

### Materials and Methods

#### Clinical datasets

Participants were recruited as a part of a prospective observational study carried out at the University of North Carolina at Chapel Hill Family Medicine Center, an academic outpatient primary care facility in Chapel Hill, North Carolina. Recruitment criteria for this study included adults 18 years of age and older receiving the seasonal trivalent inactivated influenza vaccine (TIV) for the years 2010-2011 that included the following strains: A/H1N1/California/7/2009, A/H3N2/Perth/16/2009, and B/Brisbane/60/08. Exclusion criteria included immunosuppression, immunomodulatory or immunosuppressive drugs, acute febrile illness, history of hypersensitivity to any influenza vaccine components, history of Guillan-Barre syndrome, use of theophylline preparations, or warfarin. The study cohort included both obese (body mass index bigger than 30 kg/m2, n=104) and healthy-weight (18.5 ≤BMI≤ 24.9, n=101) individuals. Patients recruited to the study provided a blood sample prior to vaccination (baseline - day 0) and one month (28-35 days) post vaccination. Blood was collected via antecubital puncture. Sera were collected using non-heparinized 10 mL vacutainers, which were allowed to clot at room temperature for 2 hours before being separated by centrifugation at 800 x g for 10 minutes. Sera were then frozen at-80oC for subsequent analysis. All procedures were approved by the Biomedical Institutional Review Board at the University of North Carolina at Chapel Hill.

We used serum samples from a prospective study of influenza vaccination conducted in 2010-11 [<u>12</u>] which included both obese (body mass index (BMI) bigger than 30 kg/m2, n=105) and healthy-weight (18.5 ≤BMI≤ 24.9, n=100) subjects. Serum samples were obtained before vaccination (baseline - day 0) and 30 days post vaccination with the 2010 trivalent inactivated seasonal influenza vaccine (TIV) that included the following strains: A/H1N1/California/7/2009, A/H3N2/Perth/16/2009, and B/Brisbane/60/08.

#### Normalization of serum concentration

Since the concentrations of different serum samples of the same individual may be different due to a variety of reasons at different time points, serum concentrations were normalized by measuring the total protein concentration in the serum using Nanodrop, and diluting all the samples to the same total protein concentration by PBS (the average concentration / 10). These diluted samples were considered as diluted 1:10, and additional dilutions were done from them.

#### Antigens

β-propiolactone (BPL)-inactivated whole influenza viruses were obtained from two resources: (1) Influenza viruses were grown in-house in embryonated chicken eggs, BPL-inactivated and purified on sucrose columns as previously described [13], and their concentrations were determined by the hemagglutinin assay, as previously described [13]. (2) Additional influenza viruses were obtained from the WHO (produced by the National Institute for Biological Standards and Control, NIBSC) as reagents for single radial diffusion (SRD) influenza potency assay with a known concentration. Recombinant HA (rHA) proteins were purchased from Sino biological (China) as purified His-tagged proteins. Most of them were produced in human HEK293 cell cultures, and some in Baculovirus-Insect Cells. Synthetic 20 amino acids (aa) peptides were synthesized at > 90% purity by CPC scientific (CA USA). Each peptide included an N-terminal KK tag as an amine group source for binding to the coated slides.

#### Antigen microarray design and spotting

To study the anti-influenza antibody repertoire we designed and spotted two types of antigen microarrays: (1) An influenza VP microarray was spotted with a panel of 34 whole inactivated influenza viruses and 23 recombinant HA proteins (Table S1) that were selected to represent the antigenic diversity of vaccine and historical human influenza strains of the H1N1, H3N2 and B subtypes from 1918 to 2016. Whole inactivated viruses were diluted in 0.005% triton and spotted at 2 HAU/μl concentration for viruses that were grown in-house in embryonated eggs, or 4 μgHA/ml for WHO viruses. Recombinant HA proteins were diluted in 0.01% triton and spotted at 16.25 μg/ml concentration. (2) A peptide microarray was spotted with 205 partially overlapping 20 aa peptides (with 15 aa overlap) spanning the full-length HA (H1) and NA (N1) proteins of the pH1N1 2009 california vaccine strain (Cal09) that was included in the TIV vaccine tested in this study. Peptides were dissolved in 20-60% dimethyl sulfoxide (DMSO) to a 2 mg/ml solution, depending on the peptide hydrophobicity, and spotted at a concentration of 1 mg/ml in 0.0025% triton X-100. Microrrays were spotted using a Scienion Sx spotter (Scienion, Germany) on Hydrogel-coated slides that bind amine groups (H slides, Schott, Germany). VP and peptide slides were printed separately, and all slides used for profiling the whole cohort were printed in a single batch to avoid potential batch effects. Each antigen was spotted in triplicate.

#### Hybridization of antigen microarrays

Human serum samples were diluted 1:3000 for measuring anti-human IgG and 1:300 for measuring anti-human IgA in a hybridization buffer that contained 1% BSA in 0.025% PBST (0.025% tween-20 in PBS). The spotted slides were blocked by 1 hr incubation on a rocker at room temperature (RT) with a chemical blocking solution (50 mM ethanolamine, 50 mM borate, pH 9.0). After blocking the slides were washed twice with 0.05% PBST, twice with PBS and once with DDW (each wash - 3 min on the rocker), and dried by centrifugation at RT for 5 minutes at 800g. Then the microarrays were hybridized with the diluted serum samples in divided trays (PepperPrint) for 2h at RT. Following washings as described above, the microarrays were incubated for 45 min with Alexa Fluor 647 labeled polyclonal anti-human IgG antibody at 1:1000 dilution (Jackson ImmunoResearch cat# 709-605-149) or Alexa Fluor 488-conjugated polyclonal anti-human IgA antibody at 1:6000 dilution (Jackson ImmunoResearch cat# 109-545-011). The secondary antibodies were diluted in 1% BSA in 0.025% PBST. To detect bound antibodies, slides were scanned on a two-laser GenePix 4400A scanner (Molecular Devices). Due to technical problems (high background or limited volume of some of the sera), a small number of the samples were not hybridized or analyzed. The analysis presented here included serum samples from 104 Ob and 99 HW individuals for peptide arrays, 94 Ob and 95 HW samples for IgG VP arrays, 87 Ob and 87 HW samples for IgA VP arrays.

#### Analysis of microarray results

Scanned slides were annotated using GenePix Pro version 7 (Molecular Devices) to obtain the mean fluorescence intensity (0 ≤MFI ≤ 65,000). The local background fluorescence intensity was subtracted from each spot MFI. The median background-subtracted MFI (median MFI-B) was selected for each triplicate of the same antigen. Data analysis was conducted using an in-house pipeline written in Python. The vaccine-induced change in the antibodies level to a given antigen was calculated as the fold-change rise from the baseline level, or as a baseline-subtracted response.

#### Magnitude and Breadth summary statistics

Normalized microarray results were analyzed using Python scripts, written by Dr. Tomer Hertz. For a given subject and a set of antigens we defined the ‘breadth’ and ‘magnitude’ as followed: (a) Magnitude - denotes the sum of median (MFI-B) results to a given set of antigens. (b) Breadth – denotes the number of antigens in a given set with a median (MFI-B) higher than a selected threshold: (MFI-B) > 2000 for peptides and recombinant HA proteins, and (MFI-B) > 4000 for whole viruses.

The magnitude and breadth were calculated for each subject at each time point and also for the post-vaccination baseline-subtracted responses, for the following sets of antigens: Cal09 H1 peptides, Cal09 N1 peptides, HA proteins of subtype B strains, HA proteins of subtype H1N1 strains, HA proteins of subtype H3N2 strains, whole viruses of subtype B strains, whole viruses of subtype H1N1 strains, and whole viruses of subtype H3N2 strains.

#### Baseline immune history (BIH) ranking

All subjects (n=205) were ranked according to the baseline magnitude or breadth to H1N1 whole viruses, or rHA proteins or H1N1 strains (rH1), or Cal09 H1 peptides, or Cal09 N1 peptides. For each ranking, the subjects were divided into quartiles, and the two extreme quartiles of highest and lowest scores, termed high-BIH and low-BIH, respectively, were compared. The number of individuals in each group varied by assay due to technical issues such as sample availability and high-background as follows:

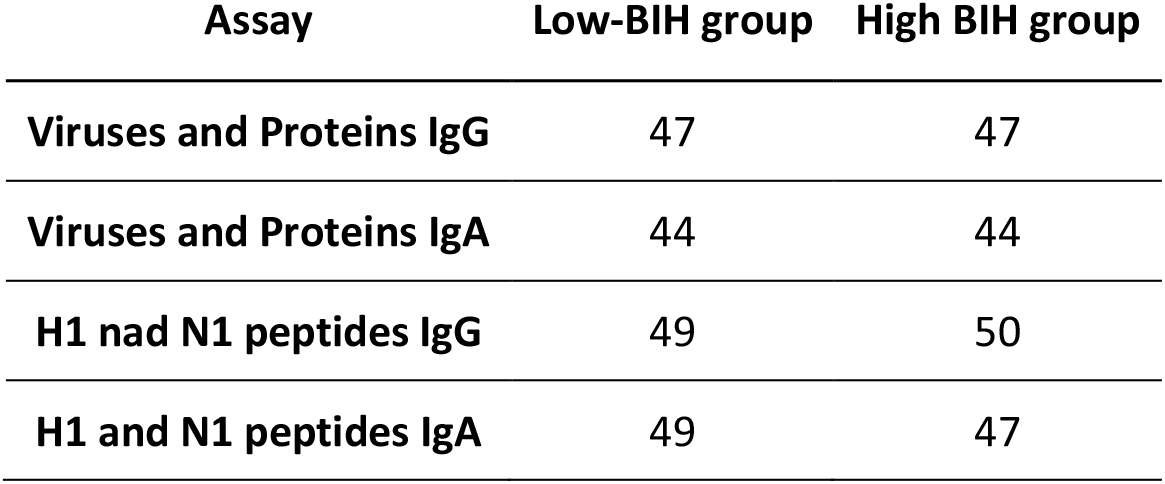

#### Vaccine responders ranking

For each individual, we computed the baseline-adjusted response to the vaccine for each peptide in the peptides microarray, by subtracting the baseline binding of antibodies to this peptide from the post-vaccination antibody binding. All subjects (n=205) were ranked according to the magnitude or breadth of baseline-adjusted responses to Cal09 H1 or N1 peptides. The subjects were divided into quartiles, and the two extreme quartiles of highest and lowest baseline-adjusted responses, termed high-responders and low-responders, respectively, were compared.

#### Statistical analysis

We used 2-sided hypothesis tests (Wilcoxon rank-sum and Fisher’s exact test) to test for differences between the distribution of breadth and magnitude scores defined above. Differences between baseline and post-vaccination responses within each group were tested using the Wilcoxon signed rank test. The relative risk (RR)and 95% CI for obese individuals or individuals <65 Y to have the magnitude of their IgG and IgA BIH magnitude of responses to whole viruses, recombinant proteins and H1 or N1 peptides to fall in the lowest quartile compared to healthy weight individuals or individuals >65 Y was calculated using the python statmoldels package. If the RR and 95% CI is smaller than 1, then bring obese or <65 y may be a protective factor, e.g., for not having a BIH magnitude in the bottom quartile. When the 95% CI includes 1, then being obese or <65 y may have no effect on the subject being in the lowest quartile of BIH response.

To predict the group of subjects based on the immune-history profiles, we used a logistic regression model and a generalized linear model (GLM) was used to predict the vaccine-induced immune responses using a logistic model for binary variables and a linear model for continuous. All models were trained using leave-one-out cross validation. Predicted values were collected over all folds for computing the area under the curve (AUC) summary stat. All continuous variables were standardized prior to training. Regularization was implemented using the elastic net package with different alpha and L1 weight parameters for each model. All models were trained using the statsmodels python package.

#### Human IgG and IgA ELISA Quantization

Commercial enzyme linked immunosorbent assay (ELISA) kits (Bethyl Laboratories, USA, cat# E80-104 for human IgG and cat# E80-1026 for human IgA) were used to quantify the total human IgG and IgA concentrations in the normalised sera, using the manufacturer’s instructions with the following modifications. The Normalized serum samples were diluted 1:24375 for human IgA ELISA and 1:243750 for human IgG ELISA. The ELISA assays were performed in 384-well white MaxiSorp Nunc plates (cat# 460372). The wells were coated with 17 µl/well of ELISA coating antibody in coating buffer, blocking and all washes were performed with 100 µl/well, and 30 µl/well diluted sera and standards were added in triplicates. Following washes, 30 µl/well HRP-conjugated detection antibody was also added. Instead of TMB, we used 30 µl/well of the SuperSignal™ West Pico PLUS Chemiluminescent Substrate (Thermo Scientific, cat# 34579, the two reagents were mixed at 1:1 ration before adding to the plate). Plates were read using a standard luminometer (TECAN infinite M200 PRO) at 600 nm. The average of each triplicate was calculated, and total antibody concentration was concluded from the standard curve.

#### Scoring HA residues based on the weights of regression model

The logistic regression model was trained based on H1/N1 peptide arrays results and assigned weights to individual features (peptides) from both the HA and NA proteins of the Cal09 vaccine strain. We used the weights from the baseline IgA or the baseline IgG models. We assigned to each residue of the HA protein the maximal weight of peptide containing the given residue. The HA residues were divided into 3 groups based on the distribution of scores of the baseline IgA or IgG.(i) Residues assigned with high positive score were associated with obesity status, (ii) Residues assigned with high negative score negative were associated with HW status and (iii) unweighted residues which had neutral or very low contribution score (below 5% of highest score in the model).

#### Curating HA domains and sites

Residues comprising the HA domain could be mapped into the following sites based on (https://www.ncbi.nlm.nih.gov/pmc/articles/PMC3410141/), The numbering is bellow is according to PDB ID : 3LZG(10.1126/science.1186430). HA1-chain A, position 11 to 325. HA2-chain B, position 1 to 175. Fusion domain: chain A, position 11 to 64, position 276 to 324. Chain B position 1 to 160. Receptor binding site (RBS)-chain A 115 to 264. Esterase domain-chain A positions 66 to 116 and 166 to 277. Conserved HA glycosylation sites - chain A positions 20-23, 33-35,289-291, Chain B 154-156. Cal09 glycosylation site - chain A positions 278 - 280. Antigenic sites - all residues comprising Cb, Sb, Sa, Ca1 and Ca2. Cb antigenic site-chain A positions 79-84. Sb antigenic site-Chain A position 187 to 199. Sa antigenic site - chain A positions 128, 129 156-161 and 162-168. Ca1 antigenic site - chain A 168-174,206-209 and 238 to 241. Ca2 antigenic site-Chain A positions 139-145 and 224-226.

#### Statistical significance test for enrichment of scored residues in functional domains

A permutation test was used to examine the significance of enrichment of scored residues in each domain. The evaluated value was different between the percentage of positively scored residues and negatively scored residues (indicating OB and HW status, respectively). When N is the number of residue in the HA protein (n =498), S is the number of residue comprising a given domain and D is the difference between positive and negative residues. The permutation test involved 10K iterations, in each iteration we sampled S residues out of N and calculated D. p value was calculated at the number of iterations where (D iteration >= D initial) divided by the number of iterations.

**Fig. S1.**
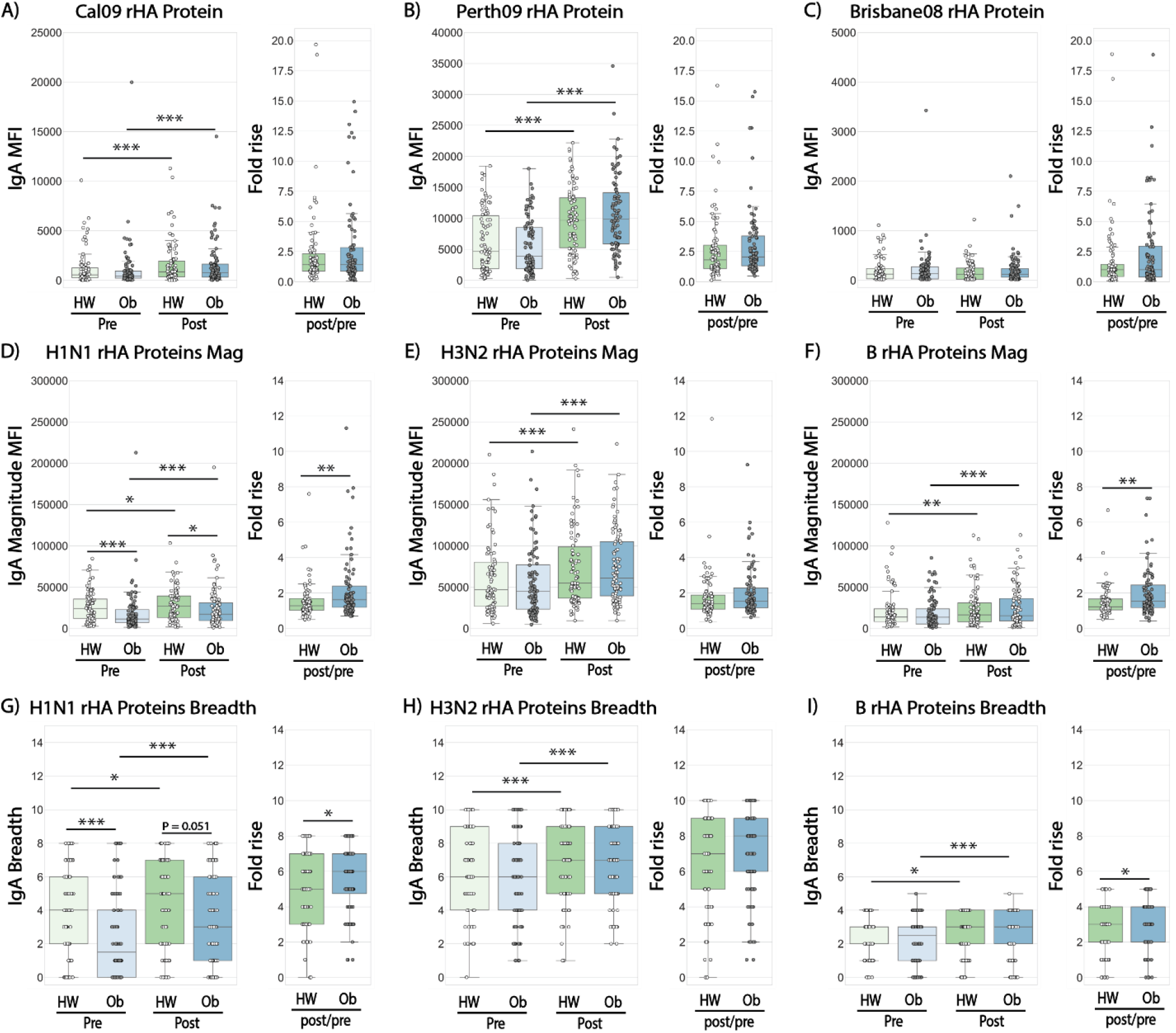
Baseline Immune History (BIH) and post vaccination IgA responses in healthy weight (HW) and obese (OB) individuals to a panel of historical influenza recombinant proteins (rHA). Baseline and post-vaccination serum samples from 82 healthy-weight (HW) and 92 obese (OB) subjects were hybridized with an antigen microarray spotted with 34 BPL-inactivated influenza viruses and 23 recombinant HA (rHA) proteins that included the three vaccine strains used in the study, for profiling of IgA binding. **(A-C)** IgA binding to the recombinant HA of H1N1 A/California/7/2009, H3N2 A/Perth/16/2009 and B/Brisbane/60/2008 vaccine strains. (**D-F**) The magnitude of IgA antibodies bound to a panel of 8 recombinant H1 (rH1) proteins (D); a panel of 10 recombinant H3 (rH3) proteins; (E) and a panel of recombinant HA antigens of 5 B strains (F). **(G-I)** The breadth of IgA antibodies bound to a panel of recombinant HA proteins, including: 8 rH1 proteins (G); 10 rH3 proteins (H); and 5 recombinant HA proteins of 5 B strains (l) proteins. The four box plots in the left portion of each panel summarize the baseline (L->R: HW: light green, Ob: light blue) and the 30-day post-vaccination (L->R: HW: dark green, Ob: dark blue) binding responses. The two boxplots on the right side of each panel represent the fold increase (L->R: HW: green, Ob: blue). Lines represent the median fluorescence intensity (MFI), the boxes denote the 25th and 75th percentiles, and the error bars represent 1.5 times the interquartile range. Statistical significance was assessed using the Wilcoxon signed rank test (pre vs. post) and the Wilcoxon rank-sum test (HW vs. Ob). * p<0.05, ** p<0.005, *** p<0.0005.

**Fig. S2.**
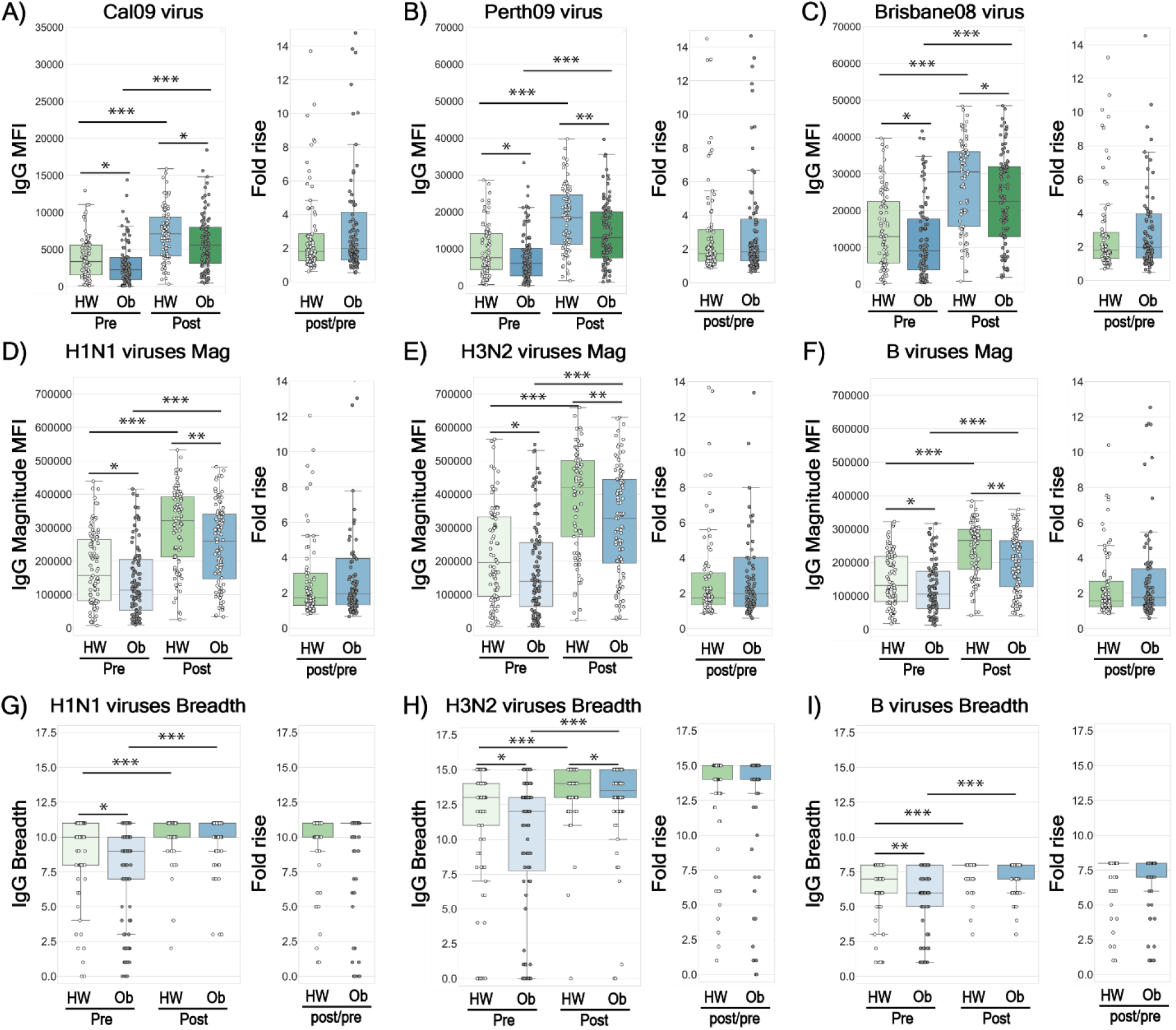
Baseline Immune History (BIH) and 30-day post vaccination IgG responses in healthy weight (HW) and obese (Ob) individuals to vaccine strains and a panel of historical influenza viruses. Baseline and post-vaccination serum samples from 89 healthy-weight (HW) and 100 obese (Ob) subjects were hybridized with an antigen microarray spotted with 34 BPL-inactivated influenza viruses that included the three vaccine strains used in the study, for profiling of IgG binding. **(A-C)** IgG binding to the H1N1 A/California/7/2009, H3N2 A/Perth/16/2009 and B/Brisbane/60/2008 BPL-inactivated viruses. **(D-F)** The magnitude of IgG antibodies bound to a panel of 11 H1N1 virus strains (D); a panel of 15 H3N2 virus strains; (E) and a panel of recombinant HA antigens of 8 B virus strains (F). **(G-I)** The breadth of IgG antibodies bound to a panel of 11 H1N1 virus strains (G); a panel of 15 H3N2 virus strains; (H) and a panel of recombinant HA antigens of 8 B virus strains (I). The four box plots in the left portion of each panel summarize the baseline (L->R: HW: light green, Ob: light blue) and the 30-day post-vaccination (L->R: HW: dark green, Ob: dark blue) binding responses. The two boxplots on the right side of each panel represent the fold increase (L->R: HW: green, Ob: blue). Lines represent the median fluorescence intensity (MFI), the boxes denote the 25th and 75th percentiles, and the error bars represent 1.5 times the interquartile range. Statistical significance was assessed using the Wilcoxon signed rank test (pre vs. post) and the Wilcoxon rank-sum test (HW vs. Ob). * p<0.05, ** p<0.005, *** p<0.0005.

**Fig. S3.**
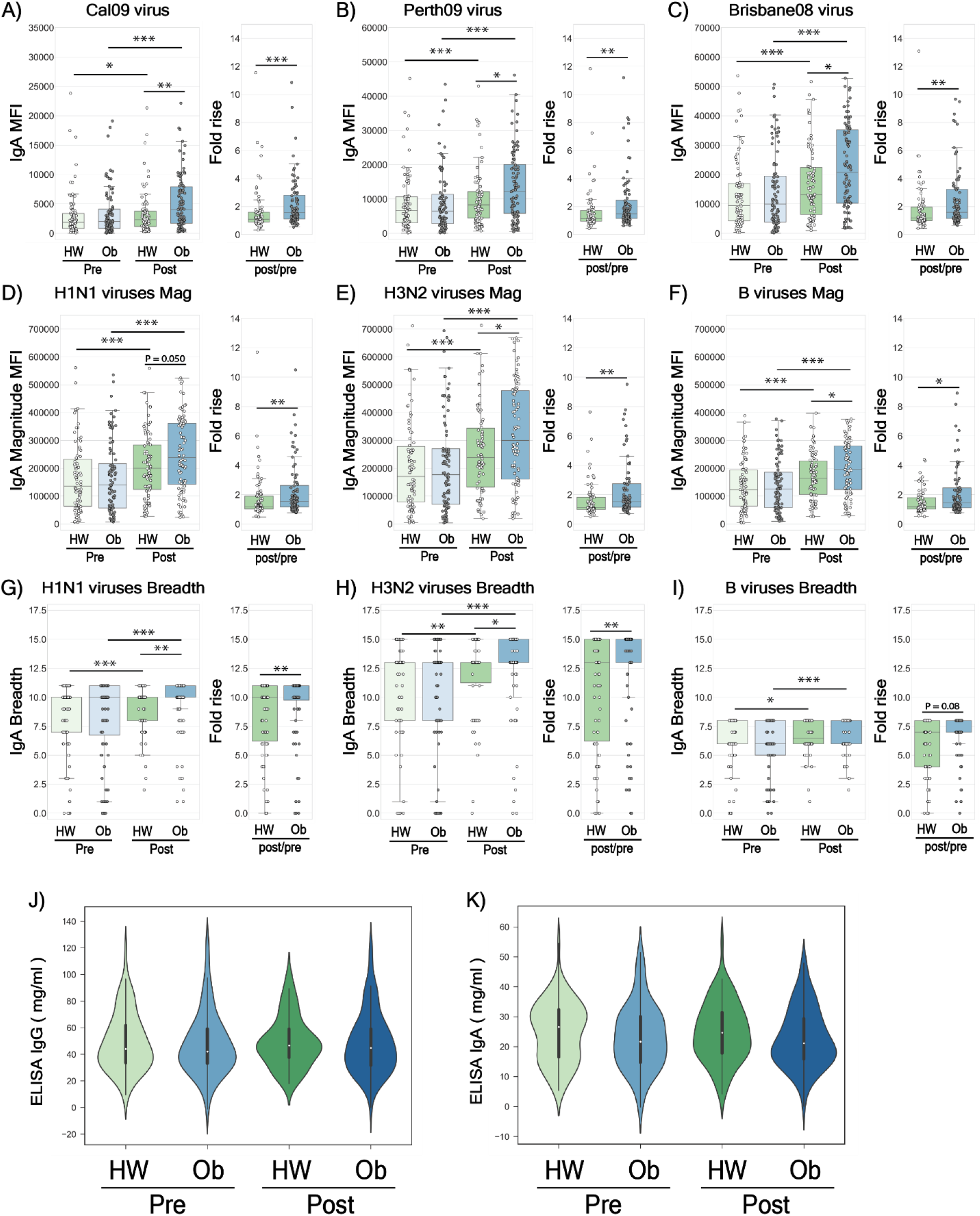
Baseline Immune History (BIH) and 30-day and post-vaccination IgA responses to a panel of BPL-inactivated influenza viruses by healthy weight and obese individuals, and Total IgG and IgA titers in the serum of obese and healthy-weight subjects. Baseline and post-vaccination serum samples from 82 healthy-weight (HW) and 92 obese (Ob) subjects were hybridized with an antigen microarray spotted with 34 BPL-inactivated influenza viruses that included the three vaccine strains used in the study, for profiling of IgA binding. **(A-C)** IgA binding to the H1N1 A/California/7/2009, H3N2 A/Perth/16/2009 and B/Brisbane/60/2008 BPL-inactivated viruses. **(D-F)** The magnitude of IgA antibodies bound to a panel of 11 H1N1 virus strains (D); a panel of 15 H3N2 virus strains; (E) and a panel of recombinant HA antigens of 8 B virus strains (F). **(G-I)** The breadth of IgA antibodies bound to a panel of 11 H1N1 virus strains (G); a panel of 15 H3N2 virus strains; (H) and a panel of recombinant HA antigens of 8 B virus strains (I). The four box plots in the left portion of each panel summarize the baseline (L->R: HW: light green, Ob: light blue) and the 30-day post-vaccination (L->R: HW: dark green, Ob: dark blue) binding responses. The two boxplots on the right side of each panel represent the fold increase (L->R: HW: green, Ob: blue). Lines represent the median fluorescence intensity (MFI), the boxes denote the 25th and 75th percentiles, and the error bars represent 1.5 times the interquartile range. Statistical significance was assessed using the Wilcoxon signed rank test (pre vs. post) and the Wilcoxon rank-sum test (HW vs. Ob). * p<0.05, ** p<0.005, *** p<0.0005. **(J-K)** Cumulative distribution plots comparing the baseline and post-vaccination total level of IgG (J) and IgA (K) in the serum samples of 205 subjects, measured by sandwich ELISA. A white dot represents the median titer.

**Fig. S4.**
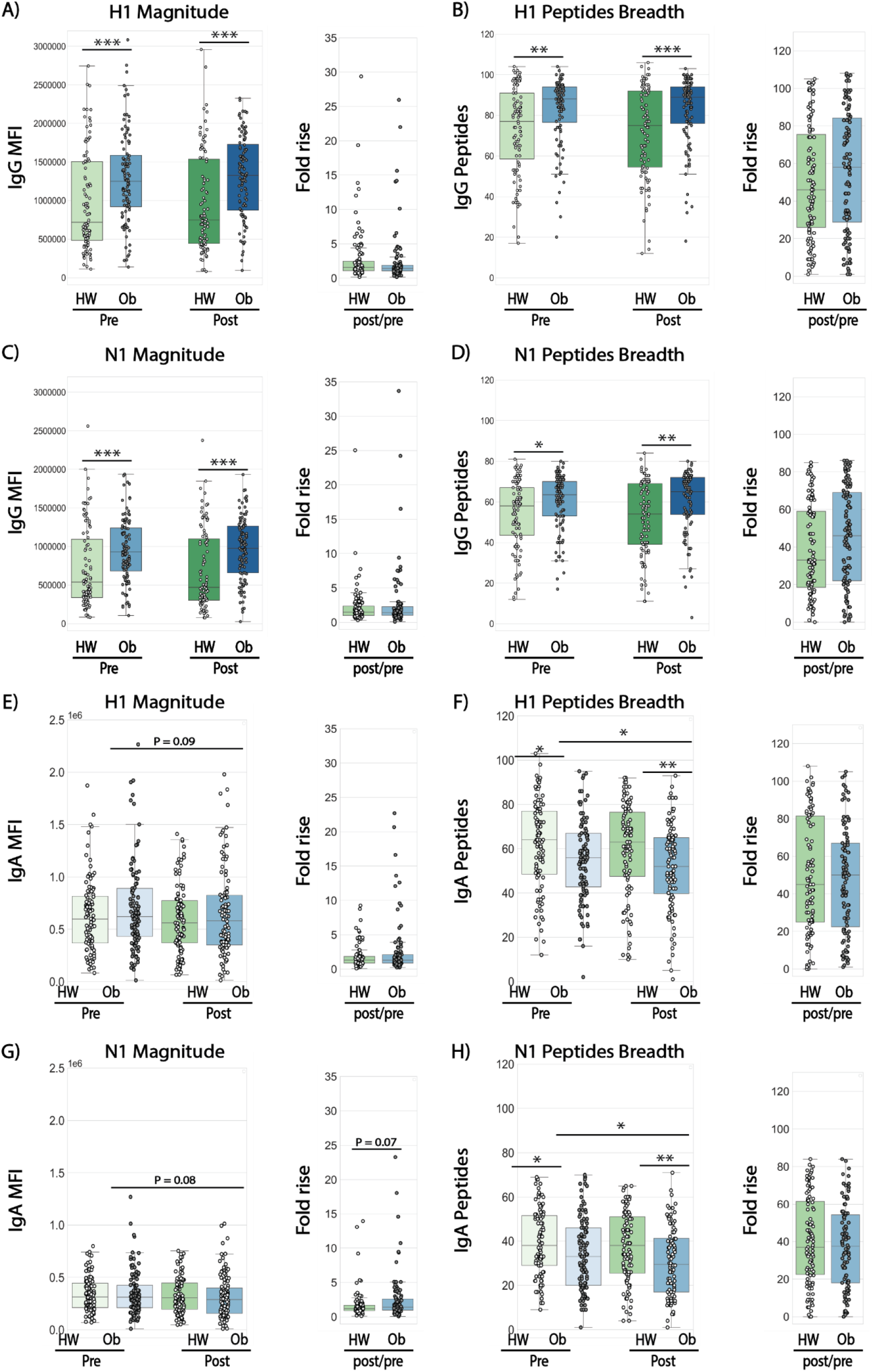
Baseline immune history (BIH), obesity-and age-associated magnitude and breadth of response profiles to influenza H1 and N1 peptides of the A/California/7/2009 vaccine strain. Arrays spotted with 20mer amino acid peptides spanning the HA and NA proteins of the H1N1 vaccine strain (15aa overlap) were used to profile the repertoires of IgG and IgA responses to linear surface epitopes of Cal09. Responses were summarized using magnitude (sum of responses to all peptides of the same protein) and breadth defined as the number of peptides from each protein to which the subject had antibodies. **(A, C)** Magnitude of IgG responses to H1 (A) and N1 (C) peptides at baseline comparing healthy-weight (HW) and obese (Ob) subjects. **(B, D)** Breadth of IgG responses to H1 (B) and N1 (D) peptides at baseline. **(E, G)** Magnitude of IgA responses to H1 (E) and N1 (G) peptides at baseline. **(F, H)** Breadth of IgA responses to H1 (F) and N1 (H) peptides at baseline. The four box plots in the left portion of each panel summarize the baseline (L->R: HW: light green, Ob: light blue) and the 30-day post-vaccination (L->R: HW: dark green, Ob: dark blue) binding responses. The two boxplots on the right side of each panel represent the fold increase (L->R: HW: green, Ob: blue). Lines represent the median fluorescence intensity (MFI), the boxes denote the 25th and 75th percentiles, and the error bars represent 1.5 times the interquartile range. Statistical significance was assessed using the Wilcoxon signed rank test (pre vs. post) and the Wilcoxon rank-sum test (HW vs. Ob). * p<0.05., ** p<0.005, *** p<0.0005.

**Fig. S5.**
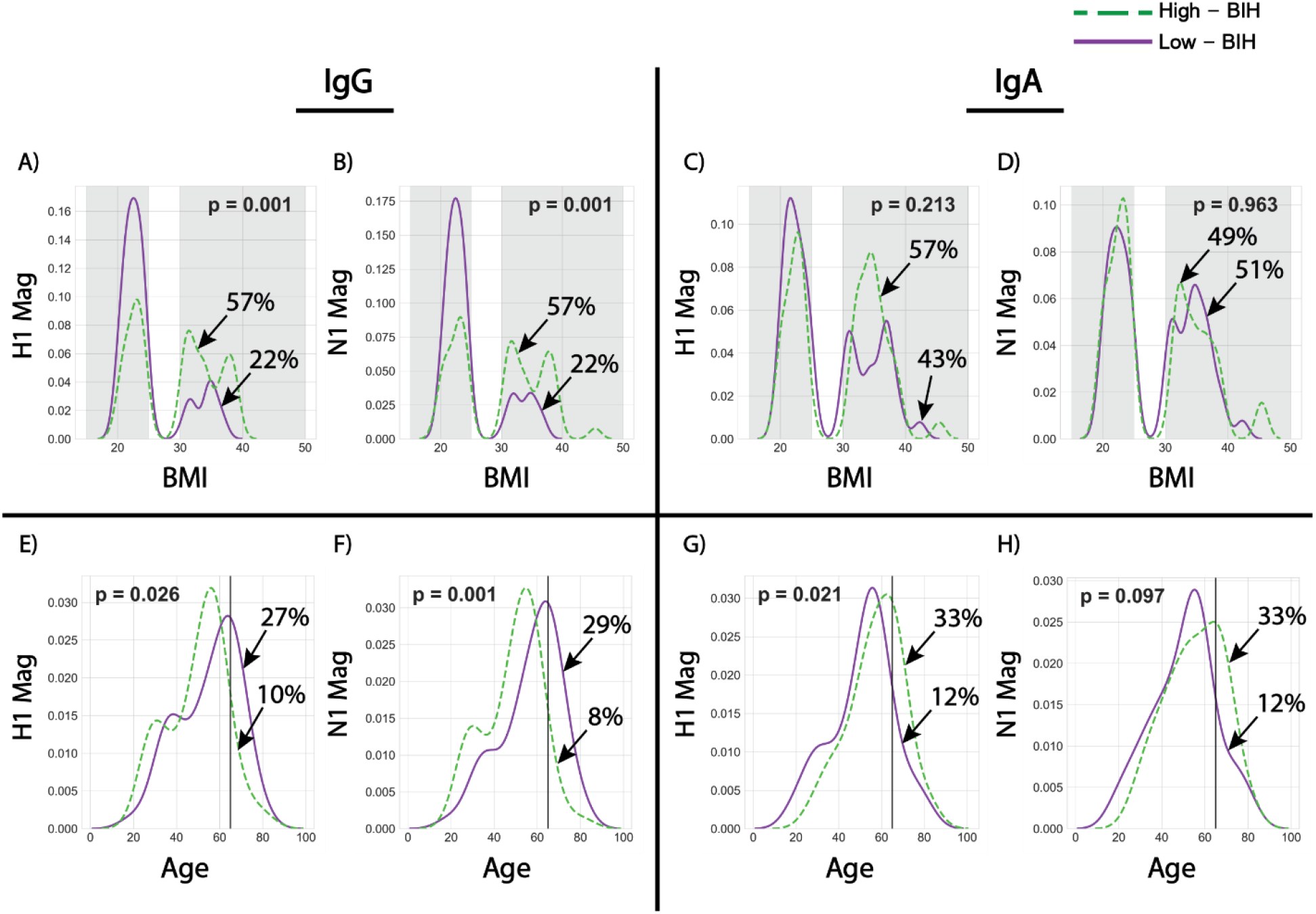
Obesity-and age-associated baseline immune history (BIH) profiles to influenza H1 and N1 proteins. (**A-D**) Distribution of the magnitude response by BMI (Panels A to D). The distributions of BMI within the low-BIH group (n=51, solid purple line) and the high-BIH group (n=51, dashed green line) were plotted for IgG and IgA responses to Cal09 H1 and N1 antigens as follows: (A) IgG magnitude against H1 peptides (B) IgG magnitude to N1 peptides; (C) IgA magnitude to H1 peptides; and (D) IgA magnitude to N1 peptides. The percentages of HW and OB subjects in the high-quartile of BIH responders are listed. Overweight subjects (25 < BMI < 30) were excluded from our analysis. The percentage of obese subjects in the low-BIH responders and high-BIH responders quartiles are listed. (**E-H**) Distribution of the magnitude of response by age. The distributions by subjects age group (<65 y and > 65 y) within the low-BIH group (n=51, solid purple lines) and the high-BIH group (n=51, dashed green lines) were plotted for their IgG and IgA responses to H1N1 antigens as follows: (E) IgG magnitude against H1 peptides; (F) IgG magnitude to N1 peptides; (G) IgA magnitude against H1 peptides; and (H) IgA magnitude to N1 peptides. The percentages of individuals > 65y in the low and high-quartile of BIH responders are listed. p values for differences between the BMI and age distributions of the low - BIH and high-BIH groups were determined using the Wilcoxon ranksum test. Obesity was associated with high IgG-BIH to peptides of the Cal09 H1 and N1 peptides. Age >65 y was associated with low IgG-and IgA-BIH to Cal09 H1 peptides, and low IgG-BIH (but not IgA-BIH) to Cal09 N1.

**Fig. S6.**
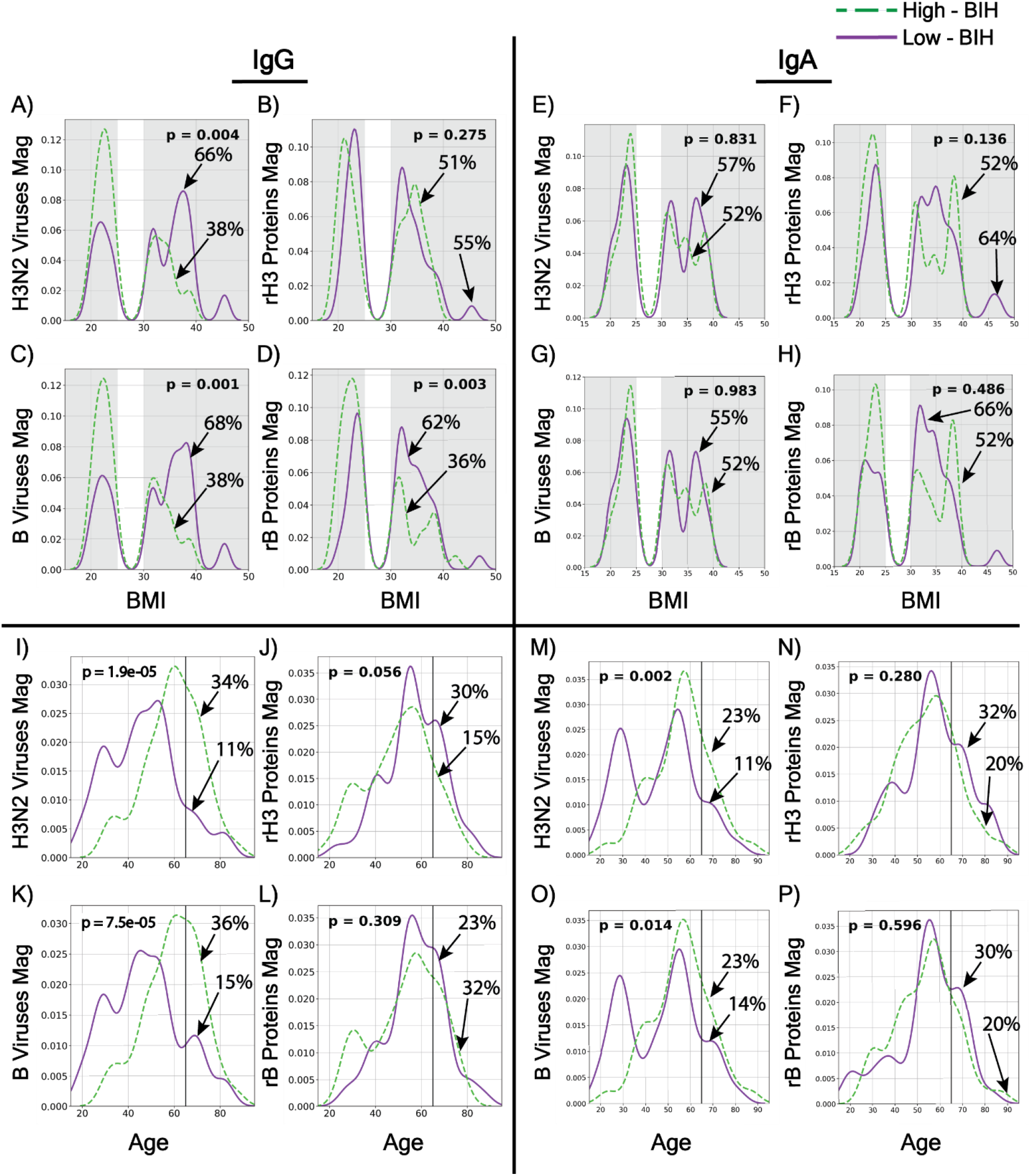
Obesity- and age-associated baseline immune history (BIH) profiles to influenza BL-inactivated H3N2 and B viruses and recombinant HA proteins. (**A-H**) Distribution by BMI. The distributions of BMI within the low-BIH group (n=47 for IgG and n=44 for IgA, solid purple line) and the high-BIH group (n=47 for IgG and n=44 for IgA, dashed green line) were plotted for IgG and IgA responses to H3N2 and B viruses and recombinant HA proteins as follows: (A) IgG magnitude against H3N2 viruses; (B) IgG Magnitude to rH3 proteins; (C) IgA magnitude to H3N2 viruses; (D) IgA magnitude to rH3 proteins; (E) IgG breadth to H3N2 viruses; (F) IgG breadth to H3 peptides; (G) IgA breadth to B viruses; and (H) IgA breadth to B proteins. (**I-P**) Distribution by age. The distributions by subjects age group (<65 y and > 65 y) within the low-BIH group (n=47 for IgG and n=44 for IgA,, solid purple line) and the high-BIH group (n=47 for IgG and n=44 for IgA,, dashed green line) were plotted for their IgG and IgA responses to H3N2 viruses and recombinant HA proteins as follows: (I) IgG magnitude against H3N2 viruses; (J) IgG Magnitude to rH3 proteins; (K) IgA magnitude to H3N2 viruses; (L) IgA magnitude to rH3 HA proteins; (M) IgG breadth to B viruses; (N) IgG breadth to rB HA proteins; (O) IgA breadth to B viruses; and (P) IgA breadth to rB HA proteins. p values for differences between the age distributions of the low-BIH and high-BIH groups were determined using the Wilcoxon ranksum test.

**Fig. S7.**
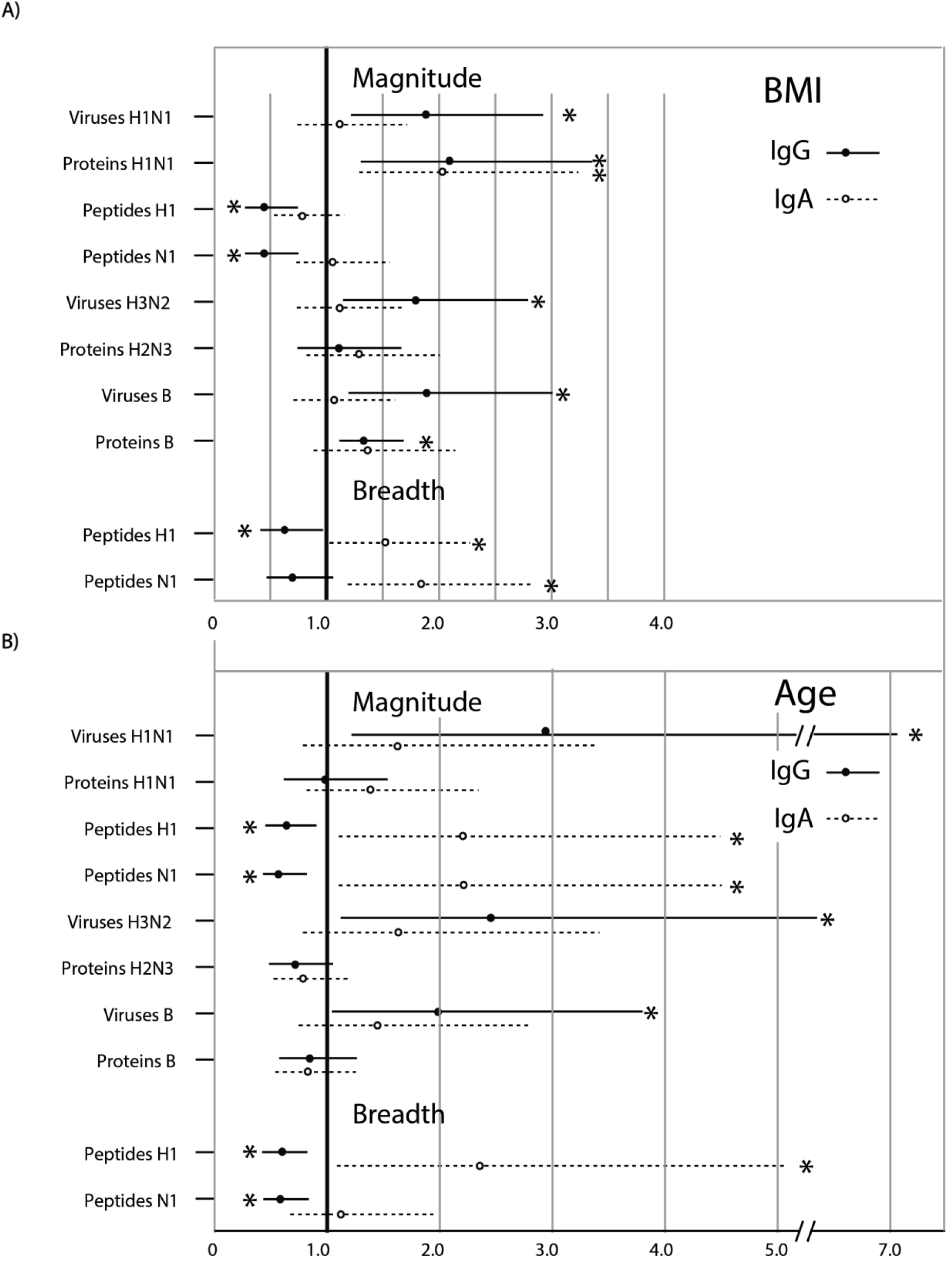
Relative risk for being in the lowest quartile of magnitude or breadth of BIH responses. **(A)** Upper panel: Relative risk (RR) for obese individuals to have the magnitude of their IgG and IgA BIH responses in the lowest quartile. The relative risk and 95% CI for obese individuals to have the magnitude of their IgG (black filled circles) and IgA (white filed circles) BIH responses in the lowest quartile compared to healthy weight individuals was calculated using an on-line calculator from MedCalk (https://www.medcalc.org/calc/relative_risk.php). The range of the 95% confidence intervals are indicated by the solid and dashed. Lines, respectibvely. **(B)** Lower Panel: Relative risk (RR) for individuals <65 y have the magnitude of their IgG and IgA BIH responses in the lowest quartile compared with individuals >65 y. The relative risk and 95% CI for individuals <65 Y to have the magnitude of their IgG (black filled circles) and IgA (white filled circles) BIH responses in the lowest quartile compared to individuals >65 Y was calculated using the Python statsmodels package. The range of the 95% confidence intervals is indicated by solid and dashed lines, respectively. The dark horizontal line indicates a RR of 1. If the RR and 95% CI is larger than 1, then being obese or <65 y may be a positive risk for having a BIH magnitude in the bottom quartile. If the RR and 95% CI is smaller than 1, then bring obese or <65 y may be a protective factor, e.g., for not having a BIH magnitude in the bottom quartile. When the 95% CI includes 1, then being obese or <65 y may have no effect on the subject being in the lowest quartile of BIH response. The specific targets are listed on the vertical axis. 95% CIs that do not include 1 are indicated by an Asterix.

**Fig. S8.**
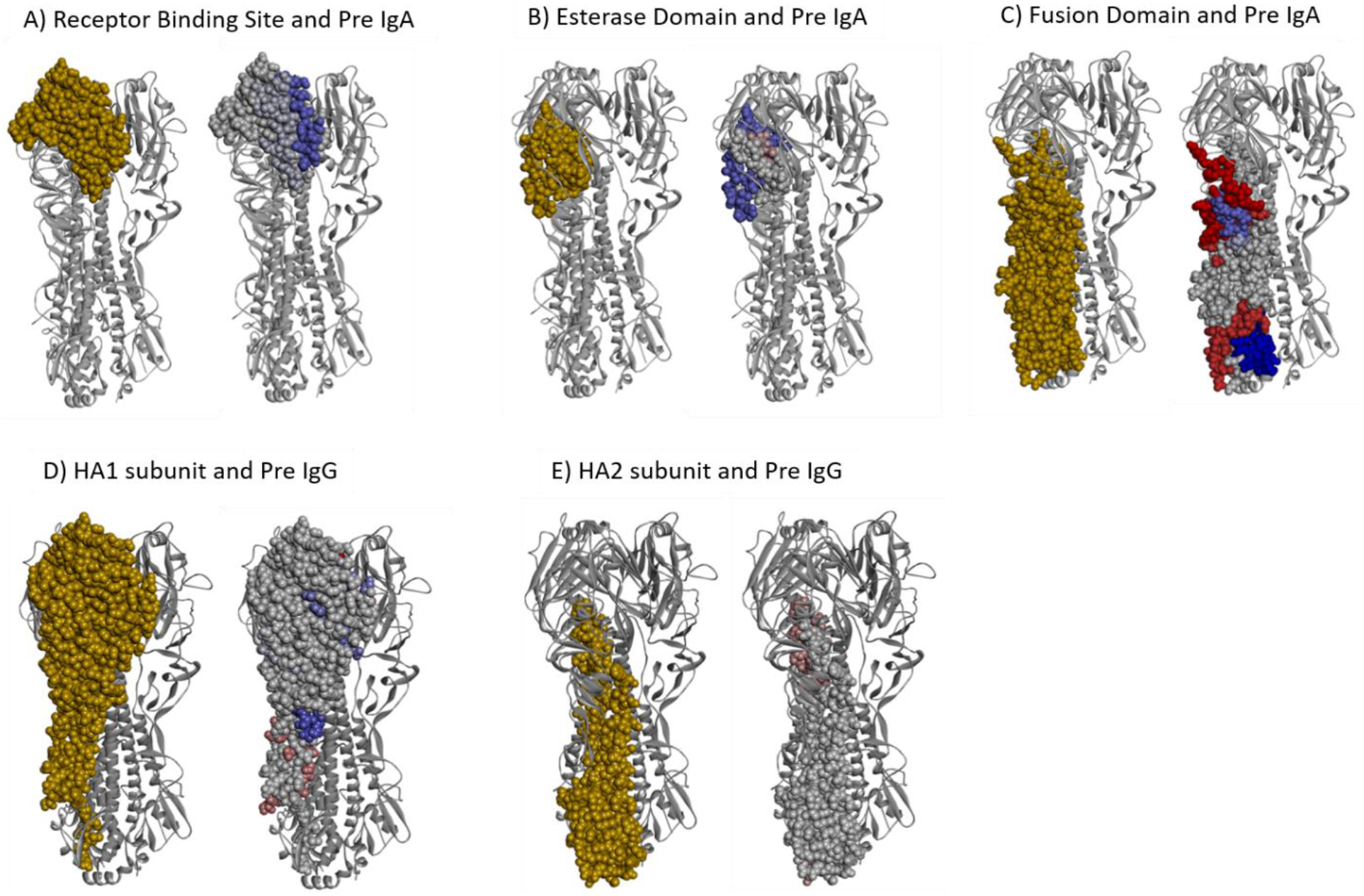
Different domains in the Cal09 HA protein are differentially targeted by BIH antibodies of obese and healthy weight subjects. A logistic regression model was trained to discriminate between HW and OB individuals using the IgG and IgA antibody profiles to the HA peptides of the Cal09 vaccine strain. The weights assigned by the model were used to score individual amino acids on the HA protein based on the maximal weight of a given position across all of the peptides in which it was included (see Methods for details). Figures were created using Discovery Studio Visualizer software (XXref) and the crystal structure of the Cal09 HA trimeric protein PDB ID: 3LZG (10.1126/science.1186430). The HA trimeric protein is presented as a gray ribbon. Residues colored in gold comprise the given site. Residues associated with HW status are colored in blue shades according to their scores. Residues associated with OB status are colored in red shades according to their scores. Sites and scored residues are presented on a single HA subunit. Left side in each panel: Dark gold spheres represent amino acid residues belonging to each of the five regions of interest mapped onto one of the three trimeric proteins: **(A)** Receptor binding site; **(B)** The esterase domain; **(C)** The fusion domain; **(D)** the HA1 subunit; and **(E)** the HA2 subunit. Right side in each Panel: Dark blue spheres represent amino acid residues within the regions of interest preferentially associated with IgG or IgA antibodies in sera from HW individuals (Panels A to D) and dark red spheres represent amino acid residues within the regions of interest preferentially associated with IgG or IgA antibodies in sera from HW individuals (Panels B, D, and E).

**Table S1.** Whole inactivated viruses and recombinant HA proteins antigens spotted on the influenza VP microarrays.

| Subtype | Strain | Vaccine Year | Whole Virus | Recombinant P<br>rotein HA |
| --- | --- | --- | --- | --- |
| H1N1 | A/WSN/1933 | - | X | X |
|  | A/Puerto Rico/8/1934 | - |  | X |
|  | A/USSR/90/1977 | - | X | X |
|  | A/Brazil/11/1978 | - | X |  |
|  | A/Chile/1/1983 | 1984-1987 | X |  |
|  | A/Singapore/6/1986 | 1987-1997 | X |  |
|  | A/Beijing/262/1995 | 1998-2000 | X | X |
|  | A/New Caledonia/20/1999 | 2000-2007 | X | X |
|  | A/Solomon Islands/3/2006 | 2007-2008 | X | X |
|  | A/Brisbane/59/2007 | 2008-2010 | X | X |
|  | A/California/7/2009 * | 2010-2016 | X | X |
|  | A/Christchurch/16/2010 | 2010-2016 | X |  |
| H3N2 | A/Bangkok/1/1979 | - | X |  |
|  | A/Leningrad/360/1986 | 1987-1988 | X |  |
|  | A/Guizhou/54/1989 | - | X | X |
|  | A/Shandong/9/1993 | - | X |  |
|  | A/Sydney/5/1997 | 1998-2000 | X | X |
|  | A/Panama/2007/1999 | 2000-2004 | X |  |
|  | A/New York/55/2004 | 2005-2006 | X | X |
|  | A/California/07/2004 | - | X |  |
|  | A/Wisconsin/67/2005 | 2006-2008 | X | X |
|  | A/Brisbane/10/2007 | 2008-2010 | X | X |
|  | A/Perth/16/2009 * | 2010-2012 | X | X |
|  | A/Victoria/210/2009 | 2011 (S) | X | X |
|  | A/Victoria/361/2011 | 2012-2013 | X | X |
|  | A/Texas/50/2012 | 2013-2015 | X | X |
|  | A/Switzerland/9715293/2013 | 2015-2016 | X | X |
| B | B/Lee/1940 | - | X |  |
|  | B/Yamagata/16/1988 | - | X | X |
|  | B/Jiangsu/10/2003 | 2005-2006 | X |  |
|  | B/Malaysia/2506/2004 | 2006-2008 | X | X |
|  | B/Florida/4/2006 | 2008-2009 | X | X |
|  | B/Brisbane/60/2008 * | 2010-2017 | X | X |
|  | B/Massachusetts/2/2012 | 2014-2015 | X |  |
|  | B/Phuket/3073/2013 | 2015-2016, | X | X |
| Number of strains: |  | 34 | 23 |  |
| Number of strains included in seasonal vaccines: |  | 25 | 18 |  |

(S) - southern hemisphere vaccine
5 \* The strains included in the vaccine given in this trial

